# Regulatory T-cells are central hubs for age-, sex- and severity-associated cellular networks during COVID-19

**DOI:** 10.1101/2022.01.06.22268711

**Authors:** Jonas N. Søndergaard, Janyerkye Tulyeu, Ryuya Edahiro, Yuya Shira, Yuta Yamaguchi, Teruaki Murakami, Takayoshi Morita, Yasuhiro Kato, Haruhiko Hirata, Yoshito Takeda, Daisuke Okuzaki, Shimon Sakaguchi, Atsushi Kumanogoh, Yukinori Okada, James B. Wing

## Abstract

Using single-cell proteomics by mass cytometry, we investigate changes to a broad selection of over 10,000,000 immune cells in a cohort of moderate, severe, and critical Japanese COVID-19 patients and healthy controls with a particular focus on regulatory T-cells (Tregs). We find significant disruption within all compartments of the immune system and the emergence of atypical CTLA-4^high^ CD4 T-cells and proliferating HLA-DR^low^CD38^high^ Tregs associated with critical patients. We also observed disrupted regulation of humoral immunity in COVID-19, with a loss of circulating T follicular regulatory T cells (Tfr) and altered T follicular helper (Tfh)/Tfr and plasma cell/Tfr ratios, all of which are significantly lower in male patients. Shifting ratios of CXCR4 and CXCR5 expression in B-cells provides further evidence of an autoimmune phenotype and dysregulated humoral immunity. These results suggest that Tregs are central to the changing cellular networks of a wide range of cells in COVID-19 and that sex specific differences to the balance of Tfr, Tfh and plasma cells may have important implications for the specificity of the humoral immune response to SARS-CoV-2.

## Main

Since the initial outbreak in late 2019 ^1^ the devastating SARS-CoV-2 pandemic and the associated COVID-19 disease have had a severe impact on the global community, making it essential that we understand this disease in greater detail. Susceptibility to infection appears to be driven by a range of factors, with risk increasing particularly with age and, to a lesser extent, male sex ^2^. Understanding the changes to the immune system of infected patients is imperative since both viral clearance and many acute symptoms are mediated by the immune system ^3, 4^. Several previous studies have analyzed the immune response during COVID-19 and demonstrated significant dysregulation of almost every immune population ^5–8^. This is also true for regulatory T-cells (Tregs), with several groups having reported some degree of disruption in their frequency, although a clear consensus has yet to emerge ^9, 10^.

Foxp3 expressing Tregs play a key role in the control of the immune system due to their ability to suppress the function of a wide range of cell types and prevent severe autoimmunity ^11^. Tregs are also known to dampen the resolution phase of an infection and have been demonstrated to have an important role in the response to various infectious diseases such as influenza and malaria ^12, 13^. Of particular relevance in the context of viral lung infections is the role of the specialized Treg subset T follicular regulatory T cells (Tfr) to control plasma cell formation, the quality of the specific antibodies, emergence of autoreactive antibodies, B-cell memory, and protection from lung damage during influenza infection ^14–16^. Several recent reports ^17, 18^ have demonstrated that many patients with COVID-19 produce autoantibodies, which may have a critical role in the progress of infection due to their ability to neutralize protective host factors such as Interferons. In some cases, these autoantibodies are a pre-existing risk factor prior to infection. However, there is also clear evidence of *de novo* generation ^19^. These factors suggest that Tregs and Tfr may be an important factor in understanding both susceptibility to, and recovery from, COVID-19.

Considering these prior findings, we hypothesized that Tregs may have potential roles both in the acute anti-viral response and the development of post-infection autoimmunity in COVID-19. In this report, we leverage the ability of single-cell proteomics (mass cytometry) to resolve rare populations, such as Treg subsets, while also retaining a broad view of the immune system in a large patient cohort. We find that subsets of Tregs are key parts of the changing cellular networks related to severity, age, and sex of patients. Most notably, we see that patients with COVID-19 have a reduced ratio of Tfr to both T follicular helper (Tfh) and antibody producing plasma cells, and that this is more severe in male patients. Our data provides cellular evidence of dysregulated antibody responses, which could explain previous reports of increased autoantibodies in male patients.

## Results

### COVID-19 generates atypical CTLA-4^high^ effector and CXCR4^high^ naïve conventional CD4+ T cells

Peripheral blood mononuclear cells (PBMCs) from 40 healthy controls and 55 COVID-19 patients (Table 1) were labelled with metal-tagged antibodies and analyzed on a Helios mass cytometer (Fig. 1). Self-organizing map (FlowSOM) clustering of CD45^+^ live cells showed clear resolution of most major immune subsets (Extended data Fig.1A). Analysis of changes to cellular frequency demonstrate that in comparison to healthy controls, severe COVID-19 patients had significantly increased frequencies of B-cells, plasma blast cells (plasma), and classical monocytes (cMono) (Extended data Fig.1B **and** 1C). In contrast, CD8 T-cells, non-classical monocytes (ncMono), conventional dendritic cells (cDCs), and plasmacytoid dendritic cells (pDCs) were all significantly reduced in a manner similar to previous reports ^7, 8^. Moderate and critical patients’ immune composition were comparable to severe at this global level with the exception that moderate patients retained a more normal proportion of CD8 T-cells (Extended data Fig.1B). Follow-up samples had mostly returned to similar proportions as healthy controls indicating that these cellular changes were transient. To obtain a fine resolution of cellular populations, we then performed subset analysis of major cell types: CD4 T-cells (CD4), CD8 T-cells (CD8), NK cells (NK), B and plasma cells (B-cells, Plasma), and myeloid cells and DCs (pDC, cDC, ncMono, cMono). Low frequencies of cell doublets and PBMC contaminating neutrophils were excluded from further analysis at this stage.

**Fig 1.**
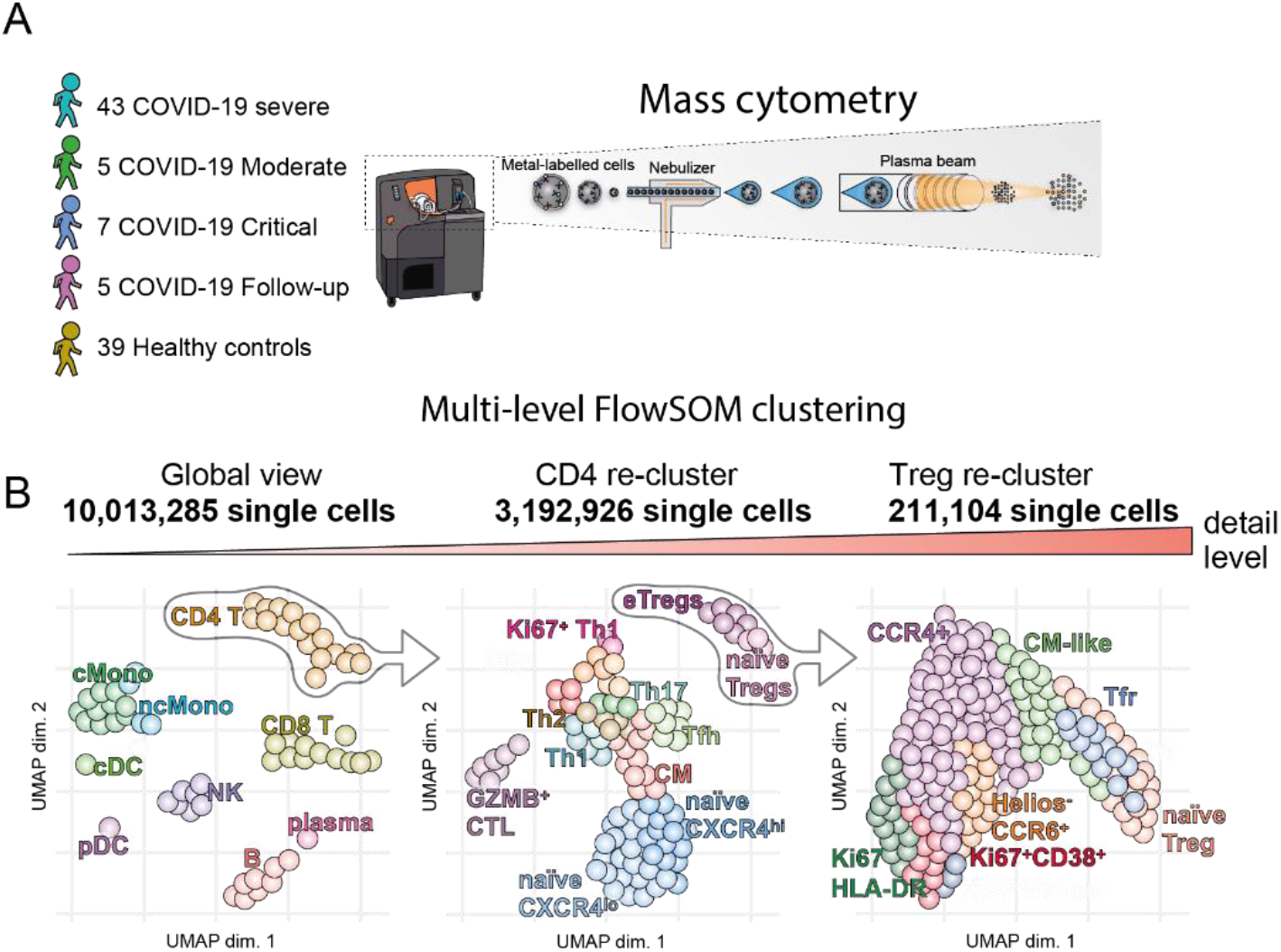
Schematic diagrams of study. **A)** Patient cohort and mass cytometry schematic. B) Multilevel FlowSOM analysis schematic.

**Table 1:** Patient characteristics

|  | Healthy controls <50 | Healthy controls 50+ | Moderate | Severe | Critical | Follow-up |
| --- | --- | --- | --- | --- | --- | --- |
| Number | 24 | 15 | 5 | 43 | 7 | 5 |
| Mean Age | 33.7 | 54.5 | 56.6 | 67.5 | 64.7 | 72 |
| Minimum age | 22 | 50 | 39 | 28 | 59 | 59 |
| Maximum age | 47 | 64 | 78 | 86 | 73 | 80 |
| Female/Total | 7/24 | 11/15 | 1/5 | 17/43 | 1/7 | 1/5 |
| Mean Days after onset | N/A | N/A | 7.6 | 10.5 | 14 | 62.4 |
| Mean Days after intubation (Intubated at time of sampling) | N/A | N/A | N/A (0/5) | 2.02 (42/43) | 3.83 (6/7) | 50.75 (2/5) |
| Mean days after starting steroids | N/A | N/A | 1.6 | 4.53 | 7.14 | 55.5 |
| Tocilizumab treated/Total | N/A | N/A | 0/5 | 2/43 | 2/7 | 0/5 |

In-depth sub-clustering of CD4 T-cells identified a range of previously described CD4 T-cell populations that we manually annotated based on examination of Uniform manifold approximation and projection (UMAP) distribution and expression of markers used for clustering (Fig. 2A). For example, central memory (CM) cells were identified as CD95^+^CCR7^+^CD45RA^−^ cells located between the naïve and effector memory areas of the UMAP, while granzyme B^+^ cytotoxic CD4 T-cells (annotated as GZMB^+^ CTL) were identified as CD95^+^CD57^+^GZMB^+^ (Fig. 2A). Changes to the proportions of clusters in COVID-19 revealed perturbations across the spectrum of naïve to effector cells (Fig. 2B, 2C). Expansion of several groups of proliferative Ki67^+^ CD4 cells was seen including a group of less differentiated Ki67^lo^ cells that retained TCF1 and CCR7 (annotated as Ki67^lo^) and two more terminally differentiated HLA-DR^+^ (HLA-DR^+^Ki67^+^) and CXCR3^+^ (Ki67^+^Th1) subgroups mostly lacking markers of stemness such as TCF1 ^20^. Non-proliferating CXCR3^+^ cells (Th1) were reduced in the COVID-19 patients, potentially as they had shifted to the Ki67^+^Th1 group (Fig. 2B). Interestingly, we observed that the Ki67^+^Th1 cluster was significantly higher in critical than severe patients (Fig. 2B, 2C) and showed extremely high levels of intracellular CTLA-4 (Fig. 2A, 2D). While CTLA-4 is not exclusively expressed by Tregs, in both healthy donor PBMCs and most highly activated environments such as tumors, effector Tregs reliably express higher levels of CTLA-4 than all other populations of effector CD4 and CD8 ^21^. However, in this case the Ki67^+^Th1 cluster had significantly higher CTLA-4 expression than even effector Tregs (Fig. 2E). To better understand the relationship between these CTLA-4^hi^ cells and other effector groups we used trajectory analysis. The COVID-19 enriched effector CD4 T-cell population had a separate trajectory from other effector CD4 T-cells, rather running in parallel to Tregs due to their relative phenotypic similarity (Extended data Fig. 2A, 2B). However, despite high CTLA-4, low levels of Foxp3 expression and intermediate levels of CD25 expression were observed in the Ki67^+^Th1 subgroup, both below the expression level of Foxp3/CD25 intermediate naïve Tregs suggesting that the Ki67^+^Th1 cluster can be considered as Foxp3^lo/–^ non-Tregs ^22^.

**Fig 2.**
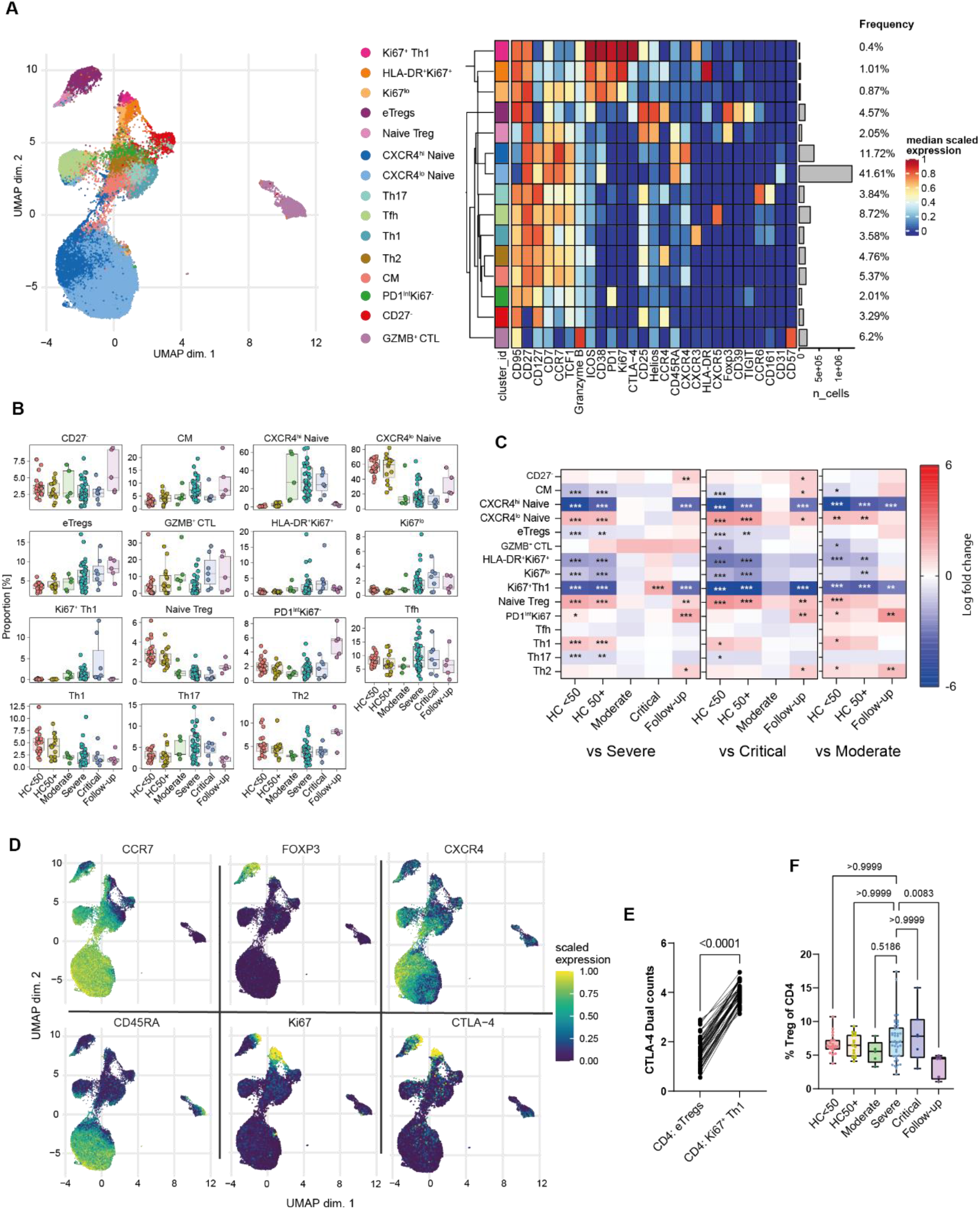
CD4 T-cell phenotypes in COVID-19. **A**) UMAP and expression heatmap of FlowSOM clusters from CD4 T-cells. The heatmap displays scaled expression of indicated markers. **B)** Frequency boxplots of proportion of CD45^+^ cell from indicated clusters. Healthy controls under 50 years of age (HC<50), healthy control of 50 or over (HC50+), moderate, severe, critical or follow-up COVID-19 patients. **C)** Comparison of the fold change (log_2_) in cluster-frequency between the indicated group and severe (left), critical (middle), and moderate (right) COVID-19 patients. **D)** UMAP feature plots of selected markers. **E)** Expression of CTLA4 on eTregs vs. Ki67^+^ Th1 cells. **F)** Percentage Tregs as a proportion of total CD4 T cells. *p≤0.05, **p≤0.01, ***p≤0.001 or value indicated on the graph. Significance by edgeR (C), Wilcoxon matched-pairs (E) or Kruskal-Wallis (F). Effector Treg (eTreg), Central memory (CM), granzyme B positive cytotoxic lymphocyte (GZMB^+^ CTL). Expression values are arcsinh-transformed (co-factor: 5) dual counts.

A significant shift in clusters within phenotypically naïve CD4 cells was also observed. This was primarily characterized by increased expression of CXCR4, the chemokine receptor for CXCL12, and a resulting shift in the frequency of clusters from CXCR4^lo^ naïve to CXCR4^hi^ naïve in moderate, severe, and critical COVID-19 patients and then returned to CXCR4^lo^ naïve in follow-up samples (Fig. 2B, 2C, 2D). These CXCR4^hi^ naïve cells retained expression of CD45RA, CD27, CD127, TCF1 and CCR7 but close examination indicated slight changes to the expression of some markers (Fig. 2A), suggesting a later stage of development. To confirm this, we verified that the CXCR4^hi^ naïve cluster had significantly reduced expression of the recent thymic emigrant marker CD31 (Extended data Fig.1C). Significant but low upregulation of CD95 was also seen (Extended data Fig.1D), suggesting a phenotypic similarity with CD95^+^CD45RA^+^ T-stem memory (TSM) cells ^23^. However, since most naïve cells in COVID-19 patients gain this phenotype, we consider it unlikely that this is driven by antigen-specific memory development but rather that CXCR4 may prime naïve T-cells for trafficking to the lungs of COVID-19 patients ^24^.

### Disruption of Tregs in COVID-19

Several reports on COVID-19 have demonstrated some degree of disruption in Tregs ^9^. We saw that Tregs as a proportion of CD4 were not changed in acute COVID-19 but appeared to be reduced at the follow-up stage (Fig. 2F). Additionally, some shift from naïve to effector Tregs was clear at the CD4 level (Fig. 2A, B). Tregs are a complex population with a range of known subtypes ^25^. To gain a better resolution of their sub-phenotypes, we performed sub-clustering of Tregs (Fig. 3A). While there is no true consensus of Treg subpopulations, we were able to recapitulate most described Treg subpopulations such as naïve, Tfr, an intermediate central memory (CM)-like population, CCR4^+^ effectors, Helios^−^CCR6^+^ cells, and several groups of highly activated HLA-DR or CD38 expressing Tregs ^22, 26–31^ (Fig. 3A, 3B). A shift towards activated Treg subtypes was seen in patients with COVID-19 with increases in the activated CCR4^+^ (annotated as CCR4^+^), Helios^−^CCR6^+^ effectors, and several groups of proliferating Tregs. (Fig. 3C, 3D). A corresponding reduction in the proportions of naïve, Tfr and CM-like was observed. The proliferating Helios^−^ cluster (Ki67^+^Helios^−^) lacked any clear association with COVID-19, while the Ki67^+^HLA-DR^+^ cluster was generally increased in all COVID-19 patient groups. However, the CD38^hi^HLA-DR ^−^ group of proliferating Tregs (Ki67^+^CD38^+^) was significantly increased in critical patients in comparison to severe or moderate groups (Fig. 3D, 3E), suggesting an association with the most severe forms of the disease.

**Fig 3.**
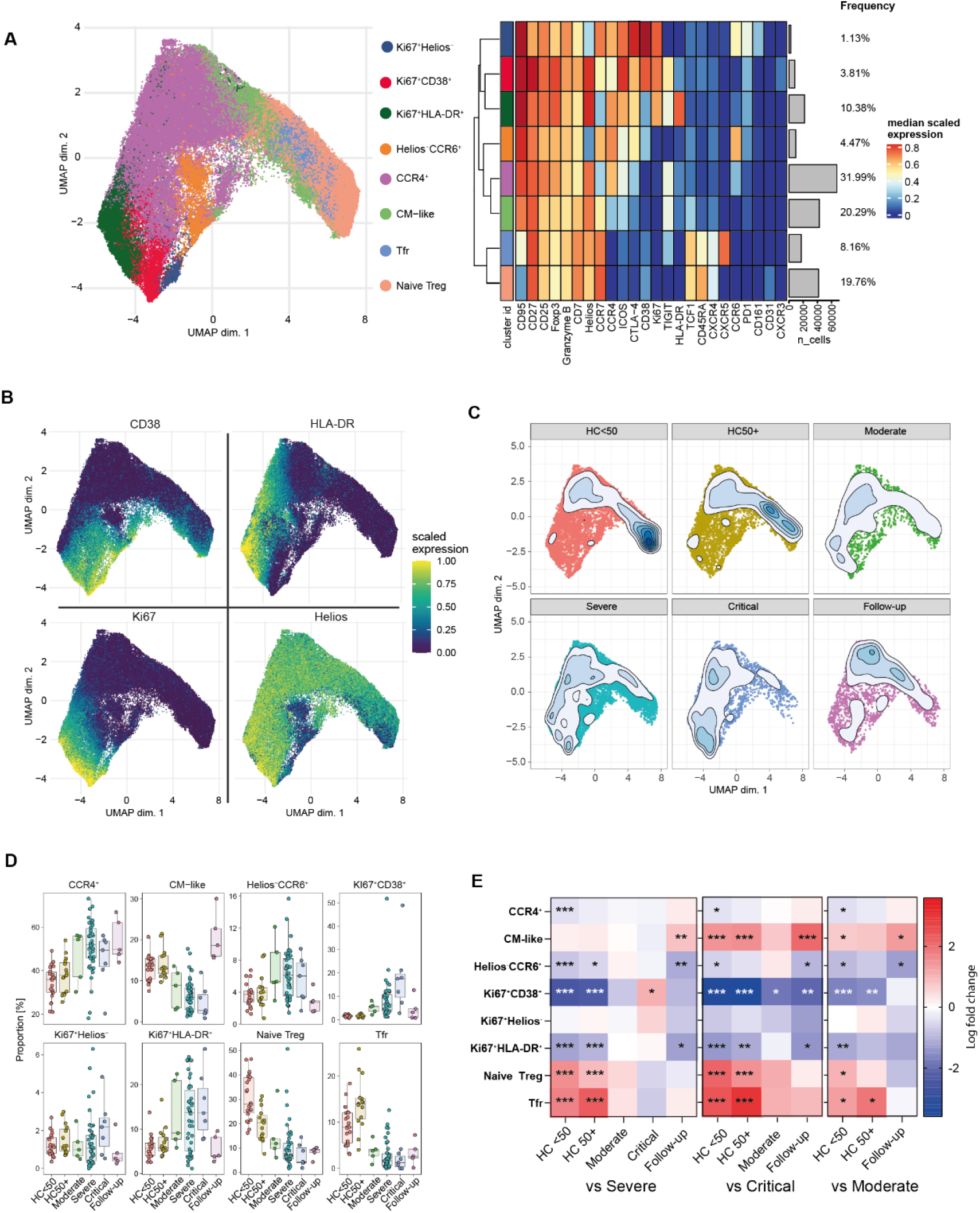
Treg phenotypes in COVID-19. **A**) UMAP and expression heatmap of FlowSOM clusters from sub-clustering of Treg cells. **B)** Scaled expression of indicated markers displayed on the UMAP. **C)** Population density of cells displayed on UMAP. **D)** Frequency boxplots of proportion of Treg cells from indicated clusters. Healthy controls under 50 years of age (HC<50), healthy control of 50 or over (HC50+), moderate, severe, critical or follow-up COVID-19 patients. **E)** Comparison of the fold change (log_2_) in Treg cluster-frequency between the indicated group and severe (left), critical (middle), and moderate (right) COVID-19 patients. *p≤0.05, **p≤0.01, ***p≤0.001 by edgeR. Expression values are arcsinh-transformed (co-factor: 5) dual counts.

### Treg subsets are central hubs in COVID-19

We also performed re-clustering of NK, CD8, B-cells, and myeloid cells and were able to replicate key findings from previous literature, demonstrating the accuracy of this analysis. Severe COVID-19 infection led to increased proportions of activated, proliferating and cytotoxic CD57^+^CD69^+^ and Ki67^+^ NK cells while Granzyme-B^lo^CD57^−^ NK and CD56^hi^CD16^lo^ NK were decreased (Extended data Fig.3A-E) ^32^. In CD8 T cells, naïve and CD161^+^CCR6^+^ mucosal associated innate T-cells (MAIT) were greatly reduced while several subgroups of proliferating Ki67^+^ effector memory like cells characterized by high expression of both CD38 and HLA-DR were increased (Extended data Fig.4A-E) ^33, 34^. Further analysis of the myeloid compartment confirmed the presence of the HLA-DR^lo^ atypical cMono and loss of intermediate and ncMono that several groups have found are associated with severe COVID-19 (Extended data Fig.5A-E) ^7, 8, 35^. The B-cell compartment was characterized by the large increase in plasma cells alongside the expansion of rare proliferating memory cells and CD11c^+^CXCR5^−^ extrafollicular B-cells similar to those observed by Woodruff *et al.* ^36^ while non-proliferating memory B-cells were generally reduced (Extended data Fig.6A-E). Since we collected information on many cell types, we next sought to use this information to determine the associations between these changing cellular populations by correlation analysis. To avoid undue influence from larger populations, such as cMono, we used subset frequencies within each level of clustering rather than as a proportion of all cells. For example, Treg: naïve is the proportion of the naïve Tregs as proportion of all Tregs. A large network of cells correlating with each other was increased in COVID-19, including groups of proliferating, or activated CD4, CD8, Treg, B-cells, plasma cells, NK (CD69^+^), and cMono. (Fig. 4A). We also visualized these changes as networks to enable a better understanding of the relationships between cells (Extended data Fig.7). Highly proliferating Tregs were seen to be in close correlation with plasma cells and HLA-DR^lo^ cMono and the Ki67^+^Th1 cluster of CD4 T-cells. Cellular groups that were decreased in COVID-19 patients included several types of less activated Tregs (Tfr, naïve and CM-like) in correlation with groups of DCs (pDC and cDC), CXCR4^lo^ naïve CD4, naïve CD8, and CD56^hi^CD16^lo^ NK cells and monocyte subgroups (intermediate, non-classical and HLA-DR^hi^ cMono). Overall, these results suggest a general shift to dysfunctional and suppressive phenotypes characterized by loss of HLA-DR on monocytes and high levels of suppressive molecules such as CD38 and CTLA-4 expressed by hyperactivated CD4, CD8 and Tregs (Fig. 4A). While this analysis revealed broad differences between healthy and COVID-19 patients, it could not clearly show differences within patient subgroups (moderate, severe, and critical patients). To further examine this, we used the same approach but excluded healthy and follow-up samples and restricted visualization of the correlation matrix (Extended data Fig.8A) to the top 5 correlations with moderate and critical patients and the level of positive correlation between these cell types. Moderate patients were associated with several populations that were also correlated with healthy controls such as naïve Tregs, CD73^+^ memory B-cells, and HLA-DR^hi^ cMono (Fig. 4B). This likely reflects that the moderate patients are at an intermediate phenotype between healthy controls and more severe patient groups. Upon examination of associations with the critical patient group, we saw that CD38^+^Ki67^+^ Tregs appears to be a central hub around which dysfunctional HLA-DR^lo^Ki67^lo^ cMono, plasma cells, CD11c^+^CXCR5^−^ B-cells, and Ki67^+^ proliferating memory B-cells were organized (Fig. 4B).

**Fig 4.**
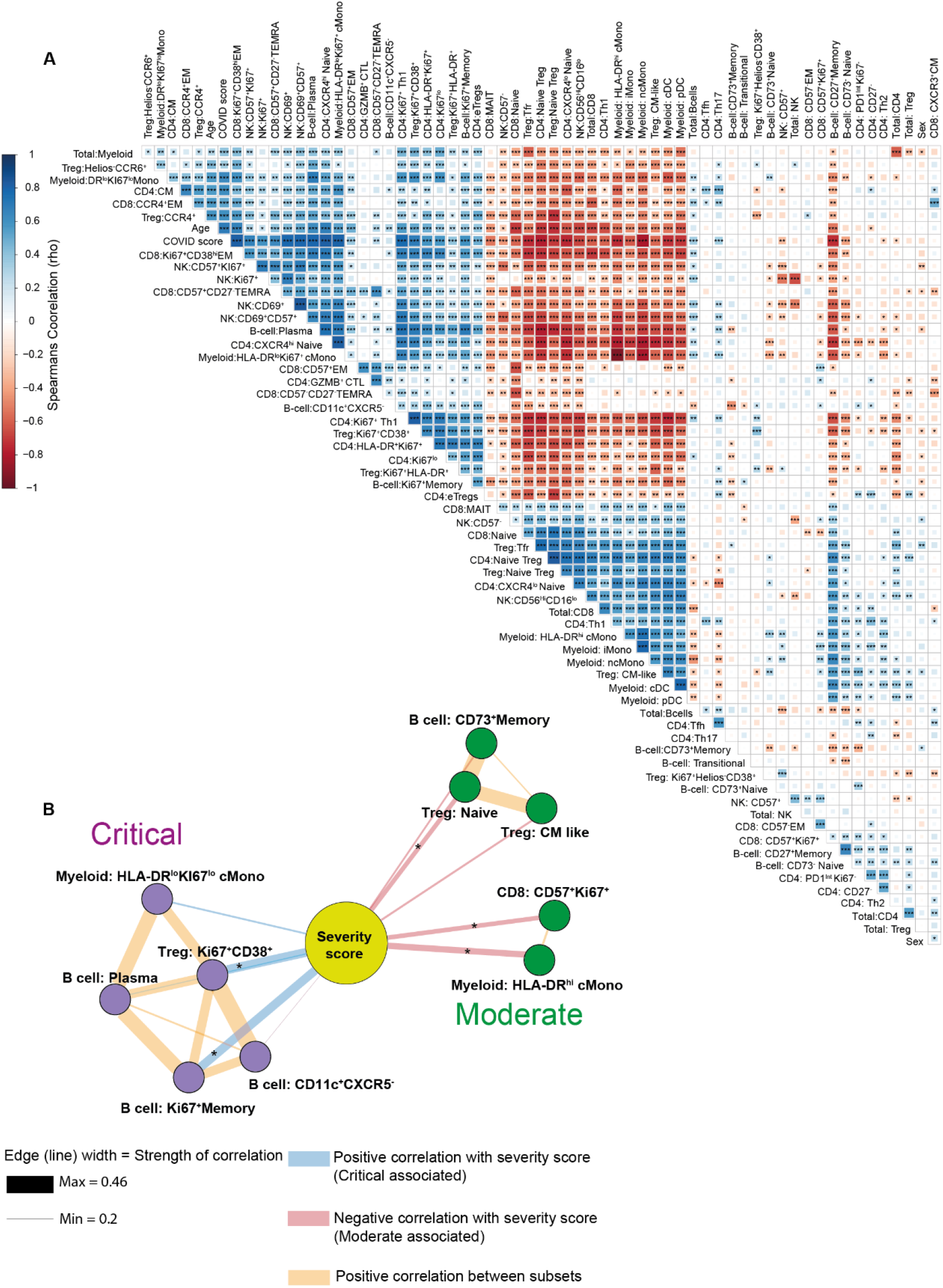
Broad changes to cellular networks in COVID-19. **A)** Spearman correlations of cellular subset frequencies between indicated subsets, COVID score (1 = healthy, 2 = COVID-19 patient) and sex (1 = male, 2= female). **B)** Top 5 correlations of subset frequencies with severity of infection scored as WHO ordinal scale (4 = moderate, 6= severe, 7 = Critical). Edge width is proportional to the Spearman correlation strength, node size is proportional to the number of connecting edges. Edges connecting to other cellular subsets are positive correlations between indicated subsets. Layout by ForceAtlas2 using edge weights as input. Significance *p≤0.05, **p≤0.01, ***p≤0.001 by Spearman (A, B). Effector memory (EM), Central memory (CM), Terminal effector CD45RA positive (TEMRA). Expression values are arcsinh-transformed (co-factor: 5) dual counts.

### Age associated changes to Tregs and CXCR4 expression

Since age is a major factor for susceptibility to COVID-19 ^2, 37^, we sought to examine the relationship between age and cellular populations. Interestingly, Helios^-^CCR6^+^ Tregs were expanded in older COVID-19 patients (Fig. 5A, 5B). In humans, Helios negativity and expression of CCR6 is characteristic of unstable Tregs capable of producing IL-17 ^28, 29^. This suggests that while Tregs are generally highly activated in COVID-19, a potentially unstable subpopulation emerges in older COVID-19 patients. The CXCR4^hi^ naïve T-cell subgroup was also increased with age in COVID-19 patients (Fig. 5A, 5C). Since we had observed increased CXCR4 expression by CD4 T cells from COVID-19 patients, we examined its expression on other cell types. In addition to the expected increase in CD4 T-cells, CXCR4 was also upregulated by CD8 T cells, B cells, cDCs, ncMono and NK cells in severe patients, and slightly decreased on cMono (Fig. 5D, 5E). These changes in CXCR4 thus appeared to be highly coordinated between cell types. The balance between CXCR4 and CXCR5 in B-cells is believed to play a key role in autoimmune disorders such as Systemic lupus erythematosus (SLE) and Rheumatoid arthritis, demonstrated by increases in a CXCR4^+^CXCR5^lo^ subset in SLE ^38–40^. We observed downregulation of CXCR5 expression by non-plasma B-cells in severe COVID-19 patients (Fig. 5F), and a greater proportion of CXCR4^+^CXCR5^lo^ B-cells, similar to those previously seen in SLE (Fig. 5G). These cells were distinct from the CD11c^+^CXCR5^−^ cells that we had found by clustering analysis, which lack CXCR4 expression (Extended data Fig.6A). The balance of CXCR5 and CXCR4 also has an important role in Tfh localization within germinal centers ^41^, and Tfh CXCR5 was reduced in severe COVID-19 patients while CXCR4 was increased (Fig. 5H). Tfh expression of CXCR4 in COVID-19 patients was also positively correlated with age (Spearman rho = 0.577, p<0.001), while CXCR5 expression was unaffected by age (rho −0.2, p=0.13).

**Fig 5.**
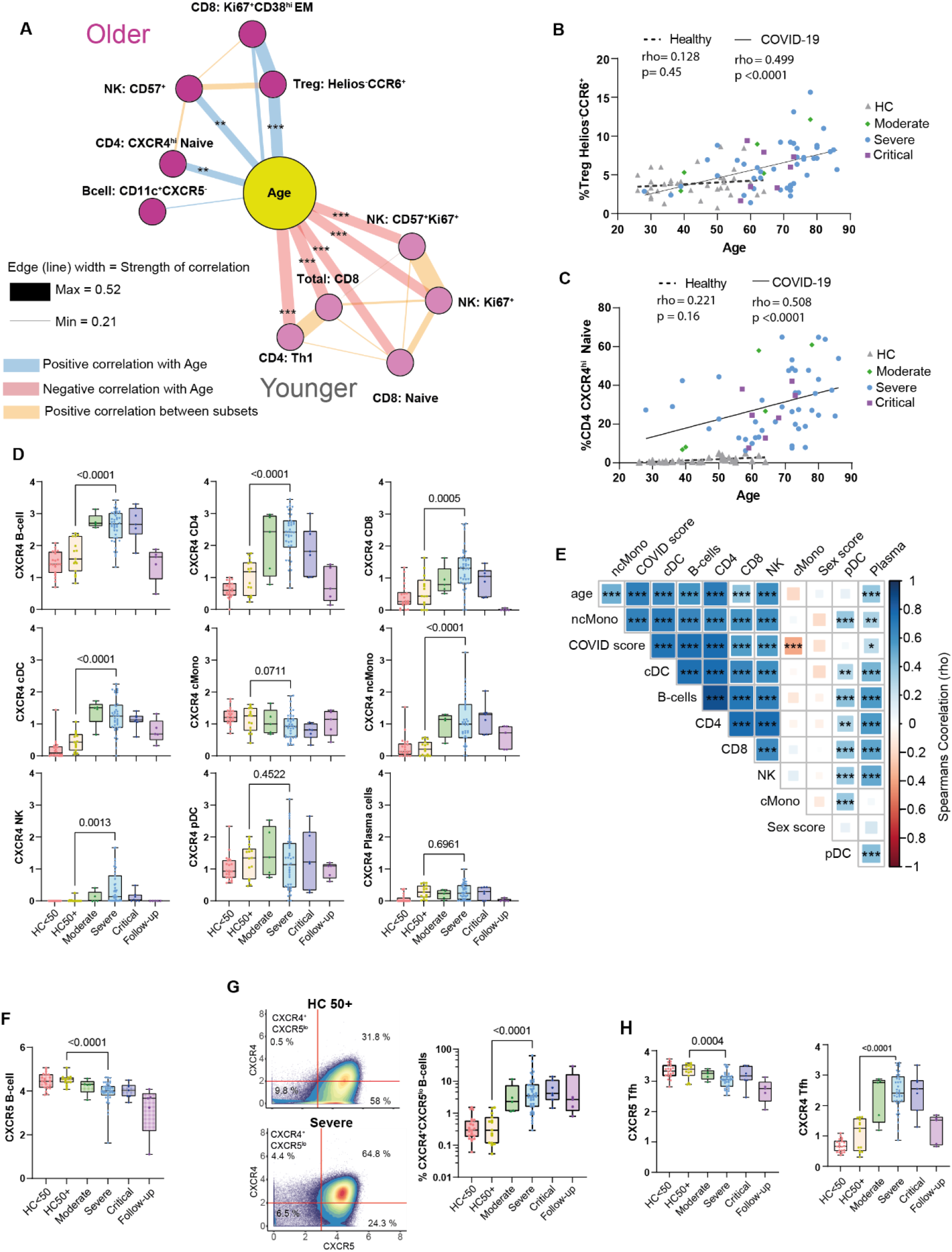
Age correlations with cellular phenotypes in COVID-19. **A)** Top 5 cellular correlations with age in severe COVID-19 patients. Edge width is proportional to correlation strength. Edges connecting to other cellular subsets are positive correlations between indicated subsets. Layout by ForceAtlas2 using edge weights as input. **B)** Correlation between age and Helios^-^CCR6^+^ Tregs. **C)** Correlation between age and CXCR4^hi^ naïve CD4 cells. **D)** Boxplots of CXCR4 expression by indicated subsets in indicated patient groups. **E)** Spearman correlations of CXCR4 expression between indicated subsets and COVID-19 status (1 = healthy, 2= COVID19) and sex (1 = male, 2= female). **F)** Boxplot of CXCR5 expression by B-cells. **G)** Gating and boxplot summary of proportions of CXCR5^lo^CXCR4^+^ B-cells. Data displayed in the gating-example is pooled from all donors of indicated groups. **H)** Boxplots of CXCR4 and CXCR5 expression by Tfh cells. Correlations in A, B, C and E Spearman rank. Significance *p≤0.05, **p≤0.01, ***p≤0.001 or indicated on graph by Spearman (B, C, E) or Mann-Whitney test (D, F, G, H). Effector memory (EM). Expression values are arcsinh-transformed (co-factor: 5) dual counts.

### Sex associated disruption of Tfr, Tfh, plasma cell ratios

Given the known importance of sex in susceptibility to COVID-19 ^2, 37^ we then sought to further dissect the associations of sex in our cohort. Since we were aware of sex bias in the critical patient group (Table 1), we restricted correlation analysis only to the severe patient group (Extended data Fig.8B) to avoid confounding effects. Female patients showed an overall increase in the proportion of B-cells (Fig. 6B), as also seen by Takahashi *et al.* ^42^. Plasma cells and Ki67^+^CD38^+^ Tregs were associated with male patients, while the top correlation with female sex was the proportion of circulating Tfr (Fig. 6A and 6B). Since a primary role of Tfr is the control of plasma cell formation ^15, 16^ this suggested a causative link between the inverse relationship of Tfr and plasma cells amongst the sexes. Upon further examination, we found that the proportion of circulating Tfr was reduced as a proportion of Tregs. Similarly, Plasma cells were also significantly reduced in females (Fig. 6B). Interestingly we also saw that healthy control females had an increased proportion of Tfr (p=0.007) compared to males and that percentages Tfr of Tregs was increased in an age-associated manner in female (rho= 0.569, p=0.01) but not male (rho=0.274, p=0.4) healthy controls. We also observed significant negative correlations between Tfr and plasma cells or Tfr and CD11c^+^CXCR5^−^ B-cells within severe patients (Extended data Fig.8B). Further division by sex demonstrated that the negative correlation of Tfr and plasma cells was significant in female but not male patients, while the Tfr/CD11c^+^CXCR5^−^ B-cell negative correlation was significant in both sexes but stronger in females (Fig. 6C).

**Fig 6.**
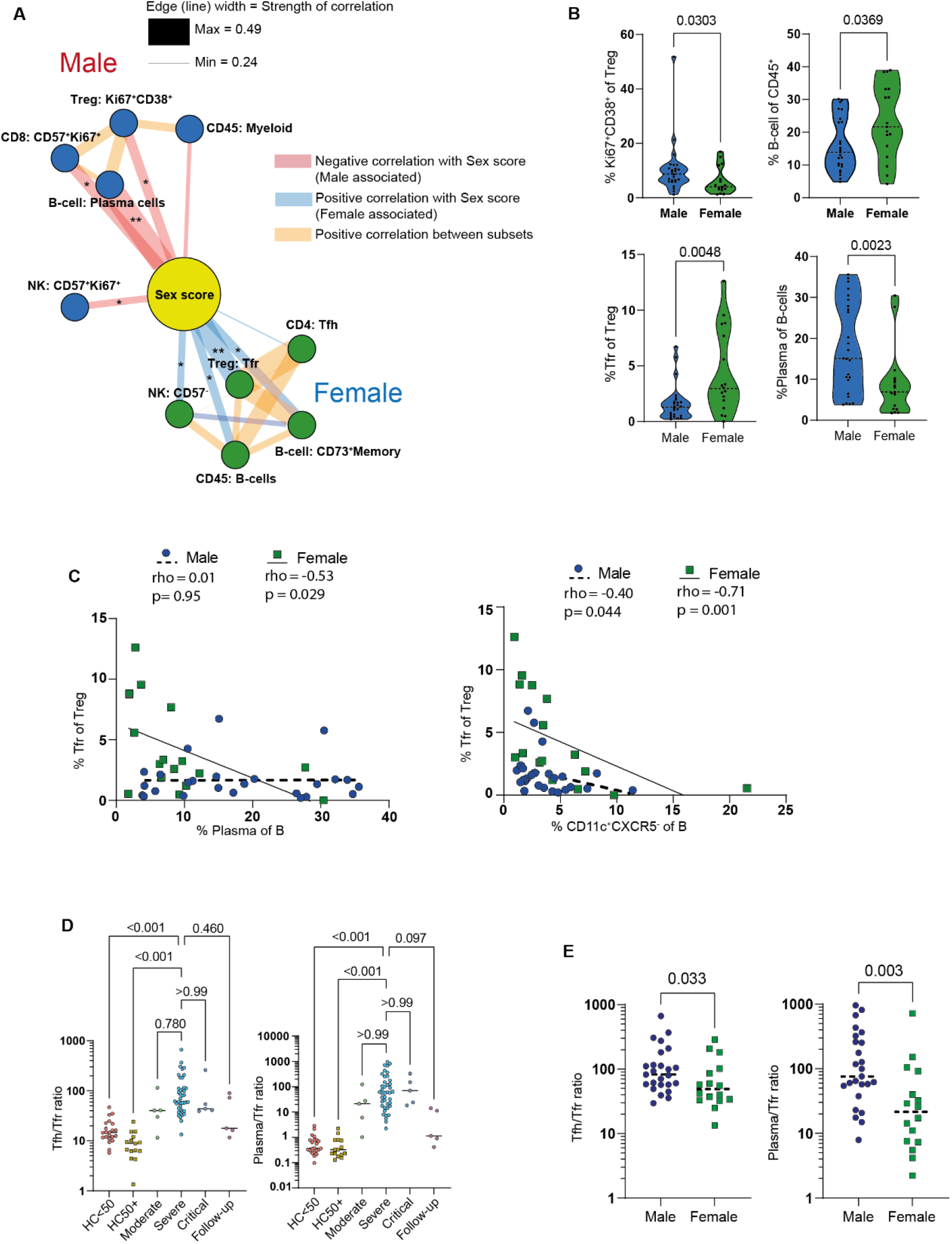
Sex correlations with cellular phenotypes in COVID-19. **A)** Top 5 cellular correlations with male and female sex in severe COVID-19 patients. Edge width is proportional to correlation strength. Edges connecting to other cellular subsets are positive correlations between indicated subsets. Layout by ForceAtlas2 using edge weights as input. **B)** Violin plots of sex specific differences of indicated cellular populations in severe COVID-19 patients. **C)** Correlation of Tfr and plasma cell frequencies in male and female severe COVID-19 patients. **D)** Tfh/Tfr ratio and plasma cell/Tfr ratio in indicated patient groups. **E)** Tfh/Tfr ratio and plasma cell/Tfr ratio in male and female severe COVID-19 patients. Correlations in A and C are Spearman rank. Significance by Mann-Whitney test (B, E), Kruskal-Wallis (E) or Spearman (C). One female patient with undetectable Tfr was excluded from the ratio analysis. Expression values are arcsinh-transformed (co-factor: 5) dual counts.

While the loss of Tfr is associated with dysregulated control of antibody responses, we also noted a positive correlation between Tfr and Tfh proportions (Fig. 6A). Previous studies have demonstrated that the balance between Tfh/Tfr and plasma cell/Tfr is more predictive of dysregulated antibody responses than their individual proportions ^27, 43^. We also examined these ratios in COVID-19 more generally and found that their disruption is apparent in almost all moderate, severe, and critical COVID-19 patients (Fig. 6D). Within severe COVID-19 patients, we also found that the ratio of both Tfh/Tfr and plasma cell/Tfr were significantly different between male and female patients (Fig. 6E). These findings suggest that disruption of Tfr function is a general factor in COVID-19 that is further exaggerated in male patients. Together with the earlier observed disruptions to CXCR4 and CXCR5 expression in B-cells this suggests a broader picture of dysregulated antibody production during COVID-19.

In summary, analysis of this cohort of COVID-19 patients has pinpointed potentially crucial roles that Tregs may play in organizing multiple aspects of the immune response in COVID-19.

## DISCUSSION

Using mass cytometry to measure broad changes to cellular phenotypes in a large COVID-19 patient cohort, we found a great number of changes, which recapitulated the findings of a number of papers, including abnormal monocytes, highly activated NK, and CD8 cells, acting as both confirmation of previous results and demonstration of the accuracy of our analysis ^7, 8, 32–35^. We also closely examined the status of Tregs during COVID-19. Several papers have observed changes to Treg during COVID-19 ^9^ variously reporting increases, decreases, or no change in Treg frequency. Alongside differences in patient cohorts, a possible cause of this variability is that not all studies use Foxp3 as part of their identification strategy and instead rely on CD127 and CD25. Since CD25 upregulation and CD127 downregulation are relatively common in highly activated non-Tregs, we would urge significant caution in reliance on surface markers alone to identify Tregs in the blood of patients with COVID-19. Additionally, Foxp3 alone is not always sufficient to correctly identify Tregs ^22^ suggesting that a range of markers and higher dimensional approaches are needed to fully separate Tregs from non-Tregs in the context of the aggressive cellular activation seen in COVID-19. In our hands, we did not see a clear change in overall Treg frequency, but in agreement with several other studies, a shift from naïve to effector Tregs was observed ^10, 44^. We confirm this broad point of a general shift in activation and extend these findings by providing further depth of analysis of Treg subpopulations. A wide range of markers have been used to define various and often overlapping definitions of effector Tregs ^25^. Here we show that while there is a general increase in activated and proliferating Treg populations, the CD38^hi^HLA-DR^lo^KI67^+^ subpopulation showed a clear stepwise increase in frequency between moderate, severe, and critical patients. CD38^lo^HLA-DR^hi^KI67^+^Tregs were also increased in all patient groups but lacked a clear association with severity. HLA-DR^hi^ Tregs are a known population with high suppressive activity ^31^ whereas CD38^hi^ Tregs are a population that have been studied most often in the context of multiple myeloma ^45, 46^. Furthermore, CD38^hi^ Tregs from either myeloma patients or healthy donors have increased suppressive function and CD38 blocking antibodies are able to reduce their function ^45, 46^. In the context of COVID-19, CD38 expression is widely induced across CD8, CD4, Treg, and plasma cells, suggesting that CD38 expression across these disparate populations may either be driven by a central factor or interaction between these cell types. In addition to Tregs themselves, we also noted an expansion of CTLA-4^hi^ proliferating T-cells, particularly in critical patients. CTLA-4 is usually expressed at higher levels in Tregs, where it suppresses the activity of T helper cells ^47^. CTLA-4 expression by non-Tregs has also been associated with exhaustion, and expression of other exhaustion associated markers such as CD38, PD1, Granzyme-B, ICOS and loss of TCF-1 ^48^. This was also the case for the CTLA-4^hi^ proliferating T-cells in the current cohort, suggesting some level of exhaustion; however, retention of CD27 and their proliferating status argues against this. Several groups have noted increased expression of CTLA-4 either in total or SARS-CoV2 specific CD4 T-cells ^5, 6^. Here we noted disruption of usual CTLA-4 expression patterns with the Ki67^+^Th1 group expressing extremely high levels of CTLA-4 expression, above that of Tregs. This high expression of CTLA-4 on proliferating T cells might, in combination with expansion of effector Tregs, be partly responsible for the establishment of a dysfunctional immune environment characterized by populations such as HLA-DR^lo^ monocytes.

Several recent studies have revealed a high frequency of autoantibodies in patients with COVID-19 ^17, 18, 49^. These may have a role both in autoimmune pathologies as well as in the resolution of infection, particularly where critical immune components such as type-1 interferon are targeted by neutralizing autoantibodies ^18, 49^. The Treg subset Tfr has been demonstrated to control autoantibody production in mice ^50^ and in humans, reduced circulating Tfr or an increased ratio of Tfh to Tfr are associated with autoantibodies and the frequency of plasma cells in patients ^51–54^. In acute COVID-19, we saw that Tfr were reduced, in keeping with another report ^55^. There is also evidence that this disruption is prolonged as COVID-19 convalescent patients have a decreased proportion of circulating Tfr and an increased proportion of activated Tfh ^56^. We found that Tfr cells were more frequent in female patients and that both the plasma cell/Tfr ratio and the Tfh/Tfr ratio was significantly higher in male COVID-19 patients. This may partly underly the finding that male patients were disproportionately over-represented amongst those with detectable autoantibodies ^18^. In the context of infection with respiratory viruses, murine models of influenza have demonstrated that without Tfr, the germinal center becomes dysregulated leading to increased proportions of antigen non-specific B-cells, increased export of plasma cells, and autoreactive antibodies ^15, 16^. Further, loss of Tfr impairs the memory response to a shared stalk domain of influenza viruses ^15^, a finding with potentially significant implications in the context of reinfection with different strains of SARS-CoV2. Tfr are suppressed by high levels of IL-2 ^16, 26^. Since plasma concentrations of IL-2 are raised in COVID-19 suggesting a role for IL-2 in the suppression of the Tfr proportion ^44, 57^. Accordingly, there is significant evidence that Tfr have an important role in suppressing plasma cell generation, improving the specificity and memory of the antibody response to acute viral infections, and preventing autoreactive antibodies from developing in this same context, all of which are of direct relevance to COVID-19. The level of interaction between Tfr and CD11c^+^CXCR5^−^ B-cells is not well established at this time; however, many Tfr are localized at the T-B border ^58^ making them well placed to prevent the initial T-cell dependent priming of extrafollicular responses and induction of B-cell class switching at this site ^59, 60^.

Further reinforcing this impression of dysregulated humoral immunity, we saw increased CXCR4 expression, a common feature of autoantibody mediated autoimmune diseases, such as SLE ^38–40^. Increased CXCR4 on B-cells or T-cells may allow them to home towards extrafollicular sites and potentially disrupt light zone organization in germinal centers ^41^. Together with increased extrafollicular CD11c^+^CXCR5^−^ B-cells, this suggests a strong influence of extrafollicular plasma cell generation in COVID-19 ^36, 61^ and a possible common link between some of the underlying mechanisms of autoantibodies in COVID-19 and SLE. The relative imbalances of Tfh/Tfr, plasma cell/Tfr, the development of B-cell phenotypes associated with SLE-like extrafollicular responses (CXCR5^−^CD11c^+^ and CXCR5^lo^CXCR4^+^ B-cells) may all contribute to the production of autoantibody responses in COVID-19 patients and requires further investigation.

CXCR4 upregulation in CD4 and CD8 cells in PBMCs from COVID-19 patients was also observed in an earlier study, although its distribution across sub-populations or other cell types was not reported ^33^. Here we find that this upregulation of CXCR4 is also generalized across many immune cell types of both myeloid and lymphoid origins, suggesting that a global factor is driving CXCR4 expression across a range of subsets in an antigen-independent manner. In addition to its role in extrafollicular B-cell responses, CXCR4 is important in the control of cellular migration to several tissues, most notably the lungs ^24^. Most immune cell types present in the bronchoalveolar lavage of COVID-19 patients express high levels of CXCR4 ^62^. CXCR4^+^CD69^+^ bystander CD4 and CD8 T-cells are seen to increase in the lungs of patients with COVID-19 and lack the tissue-resident marker CD103, suggesting recent infiltration from the periphery ^63^. We speculate that the CXCR4^hi^ naïve CD4 cells in the blood observed here, which lack CD69 expression, may be primed for non-antigen specific infiltration into the lungs and their increased frequency in older COVID-19 patients may play a role in their greater susceptibility to disease. Interestingly, CXCR4 expressing naïve CD4 T-cells are also induced in a murine model of polymicrobial sepsis, and mortality is significantly reduced by blockade of CXCR4 in that setting ^64^. Furthermore, CXCR4 signaling blocks production of IFN-I from influenza infected PBMCs, PBMCs from SLE patients, and PBMCs from COVID-19 patients ^65–67^. These findings collectively make the case that dysfunctional cellular migration, inflammation, and autoantibody production in COVID-19 may be targetable via CXCR4. Steroids are also known to induce both CXCR4 expression and changes to CXCR4 signaling via Lck ^68^. Since a high proportion of COVID-19 patients, and all patients in this study, are treated with steroids, some consideration of the effect of these drugs on CXCR4 and the relative importance of CXCR4 expression to the efficacy of these drugs in the context of COVID-19 patients is required.

In summation, in all cases, Treg subgroups were central parts of the top 5 most severity-associated (Ki67^+^CD38^+^: critical COVID-19, naïve Tregs and CM-like Treg: moderate COVID-19), age-associated (Helios^-^CCR6^+^: older patients) or sex-associated (Tfr: female patients, Ki67^+^CD38^+^:male patients) cellular populations in COVID-19 and occurs in close coordination with other described such as cMono dysfunction, NK cell activation, high proportions of plasma cells and increased extrafollicular B cells. While further work is required to truly separate cause from effect, the reduction of Tfr in all COVID-19 patients, that is further exaggerated in male patients, may underly dysregulated antibody production. While widespread induction of CXCR4 expression may also have an important role in the balance of extrafollicular antibody responses and trafficking of cells to the lungs.

## MATERIALS AND METHODS

### Study design

PBMC samples were collected from a cohort of COVID-19 patients and healthy controls (Table 1). We enrolled hospitalized cases diagnosed as COVID-19 by physicians using clinical manifestation and PCR test results. Samples were collected from August 2020 to May 2021 at Osaka University Hospital. Control subjects were collected at Osaka University Graduate School of Medicine and affiliated institutes. Due to their generally lower age the healthy control (HC) group was split by age groups into those above (HC 50+) and below 50 (HC <50) years of age to allow closer comparison of COVID-19 patients with similarly age-matched controls. Patients with COVID-19 were grouped by the WHO eight-point ordinal scale for clinical improvement ^69^, 4 = Moderate (oxygen by mask or nasal prongs), 6 = Severe (intubation and mechanical ventilation), 7 = Critical (ventilation + additional organ support – pressors, RRT, ECMO). All participants provided written informed consent as approved by the ethical committees of Osaka University Graduate School of Medicine, and affiliated institutes.

### Mass cytometry antibody production

Indium-113 and −115, and Gadolinium-157 were obtained from Trace Sciences, cisplatin-195 and −196 were obtained from BuyIsotope. Indium and lanthanide isotopes were conjugated to antibodies with the MaxPar conjugation kit (X8 polymer), while Cadmium isotopes were conjugated to antibodies with the MaxPar conjugation kit (MCP9 polymer) according to the manufacturer’s instructions. Platinum-labelled antibodies were conjugated with cisplatin as previously described ^70^. Conjugated antibodies were stored in PBS-based antibody stabilizer or HRP-protector stabilizer for cadmium labelling (Candor Biosciences). All antibodies were titrated for optimal staining concentrations using control PBMCs.

### CD45 barcoding and cell staining

A total of 9 separate experiments were performed. In each experiment, up to 1.5 × 10^6^ cells per sample were labeled with a six choose-two pattern of anti-CD45 barcodes (113In, 115In, 194Pt, 195Pt, 196Pt and 198Pt) to give a combination of up to 15 barcoded samples per experiment. Samples were incubated with CD45 barcodes together with FC-block and anti-CXCR5 biotin (Table S1A) for 30 min at room temperature (RT), and then washed twice in CyFACS buffer (PBS with 0.1% BSA and 2 mM EDTA). Barcoded cells were then pooled and washed once more with CyFACS buffer. Then, cells were stained with a metal-conjugated surface stain antibody cocktail for 45 min at RT (Table S1B). Cells were then washed twice in CyFACS buffer, stained for viability with dichloro-(ethylenediamine) palladium (II) (DCED palladium, (Table S1C) ^71^ in PBS for 5 min at RT washed and then fixed and permeabilized using the Foxp3 Transcription Factor Staining Buffer Set according to the manufacturer’s protocol (eBioscience). Cells were subsequently stained with a metal-conjugated intracellular antibody cocktail for 45 min at 4 °C (Table S1D) and then washed twice in CyFACS buffer and once in PBS. Cells were then fixed overnight in 1.6% formaldehyde solution containing DNA Cell-ID Intercalator-103Rh (Fluidigm). While DCED palladium contains approximately 11% 110Pd, we find this does not adversely affect resolution of 110Cd based CD3 staining in live cells.

### Mass cytometry data acquisition

Prior to data acquisition, cells were washed once in CyFACS buffer and twice in MilliQ H_2_O. Barcoded samples were split into separate tubes of up to 2×10^6^ cells, and centrifuged, the supernatant was removed, and samples were left as pellets until shortly before running each tube. Cells were then diluted to 1×10^6^ cells/mL in Milli-Q H_2_O containing 15% EQ Four Element Calibration Beads (Fluidigm) and passed through a 35μm filter immediately before running. Cells were acquired at a rate of 200 to 300 cells/s using a Helios mass cytometer (Fluidigm). Flow Cytometry Standard (FCS) files were normalized to EQ bead signals by the Fluidigm normalizer software.

### Mass cytometry data analysis

For analysis of the mass cytometry results, gating and de-barcoding was performed manually using Cytobank software (Beckman Coulter). Cells were initially gated as live, DNA^+^, CD45^+^ singlets with normal ion cloud Gaussian parameters. Nine Batch control samples (using two lots of heathy PBMCs) were examined for signs of batch effect and then excluded from further analysis. Three severe COVID-19 samples and two healthy control samples with low cell recovery <10000 and a high proportion (>10%) of dead cells by palladium inclusion were removed from the analysis. All other samples had a viability of >90% prior to the removal of dead cells by gating. A maximum of 200,000 cells per sample was used for analysis. Following these data filtering steps, a median of 100,943 (minimum of 18,000) cells per sample were used with a total dataset size of 10,013,285 live CD45^+^ singlets (Fig. 1A).

All dual count data channels were arcsinh-transformed (co-factor: 5) then compensated by the CATALYST R package preprocessing workflow (1.14.0) ^72^ in R (4.0.3). Analysis of data was primarily performed as in “CyTOF workflow: differential discovery in high-throughput high-dimensional cytometry datasets” version 4 ^73^ as implemented in the CATALYST R package (1.14.0) with packages cowplot (v1.1.1), flowCore (2.2.0), diffcyt (1.10.0), scater (1.18.3), SingleCellExperiment (1.12.0), ggplot2 (3.3.3). All cells were clustered by FlowSOM in the CATALYST R package with both x-dim and y-dim set to 10 to provide 100 initial SOM clusters and the consensus meta-clustering level varying from 50 to 20 in line with the expected complexity of the population. The initial 100 SOM clusters and meta-clustering were then examined manually (by expression heatmaps and UMAP or t-SNE) to find the point at which significant populations of interest were inappropriately merged. The meta-clustering level above this point was selected and used as the basis for manually merging populations to annotated subpopulations with clearer interpretations or dynamics. In all cases, initial analysis was rerun several times with new seeds to confirm that similar populations were being reproducibly found before proceeding to the refinement of the cluster numbers. For in-depth analysis of subpopulations, all cells of a particular group of interest (such as CD4 T-cells) from the CD45^+^ dataset were selected and separately processed by the same analysis workflow. CD8 cells were subject to a further round of filtering using TCRα/β and CD56 to separate them from TCRα/β^−^ or CD56^+^ cells presumed to be gamma delta T-cells and NKT that initially clustered together with CD8 in the first round of analysis. For each analysis of separate subpopulations (CD4, CD8, B-cells etc.) markers used as the basis for clustering “type markers” ^73^ were altered to select for those with a clear dynamic range of expression or biological interpretability and maximize signal to noise by exclusion of irrelevant markers. All “type markers” are displayed in expression heatmaps (Fig. 2A, 3A, **S1A, S3A, S4A, S5A and S6A**). Markers displayed in expression heatmaps were trimmed to the 99% percentile of each marker, scaled and then aggregated, preserving both information about expression differences between markers and between clusters ^73^. For dimensionality reduction samples were down-sampled to a maximum of 1000 cells per sample. UMAP was performed with nearest neighbors set to 15 with the exceptions of the CD8 and NK UMAPs which were set to 20 and 25 respectively for clearer plotting. Markers displayed in UMAPs were trimmed to the 99% percentile and then scaled. Contours were added by ggplot2 (3.3.3) and RColorBrewer (1.1-2). Differential cluster abundance analysis by edgeR was performed with diffcyt (v1.10.0) ^74^ as implemented in the CATALYST R package (v1.14.0). Wilcoxon matched-pairs, Mann Whitney or Kruskal-Wallis tests were performed in GraphPad prism (9.2). Fold change heatmaps were made in GraphPad prism with output from EdgeR. In all cases expression values are derived from arcsinh-transformed (co-factor: 5) dual counts. Except when indicated all samples were used in all analysis to give the following n numbers. HC<50 n=24, HC50+ n=15, Moderate n=5, Severe n=43, Critical n=7, follow-up n=5.

### Trajectory analysis

Trajectory analysis was done with PAGA tree in dynverse ^75, 76^ with packages dynwrap (1.2.1), dynplot (1.0.2.9000), dynmethods (1.0.5), dynguidelines (1.0.1), and dynfeature (1.0.0.9000) in R (4.0.3). Prior to analysis, samples were arcsinh-transformed (co-factor: 5), down-sampled to 500 cells per sample, and trajectory starting point defined to be in the recent thymic emigrant naïve CD4+ T cell area (CD45RA^hi^, CD31^hi^). All features except lineage markers (namely CD11b, CD3, CD4, CD8, CD14, CD11c, TCR α/β, CD16, CD56, CD19) were used for the trajectory analysis.

### Correlation analysis

Correlation analysis was performed using package corrplot (0.84) and readxl (1.3.1) in R (4.0.3). Correlation analysis used pairwise spearman rank correlation. Significance analysis of correlations used two-sided Spearman rank. Line graphs and Spearman correlation analysis in Fig. 5B, 5C **and** 6C were performed in GraphPad prism (9.2). Lines are linear correlation.

### Network diagrams

Network diagrams of correlations were produced in Gephi software (0.9.2). For the purposes of easier visual display, the negative correlations to the central nodes (COVID score, age, or sex score) were converted to positive values (x*-1). Only positive correlations between cell types were retained. In Fig. 4B, 5A, 6A an ego network with a depth of one was used to display populations with a direct connection to the central node. The top 5 positive and negative correlations with the central node are displayed. Layout was performed with ForceAtlas2 ^77^ with the following settings. Tolerance 1, scaling 25, gravity 1, prevent overlap ON, edge weigh influence 1. Following layout Edge widths were rescaled to a minimum of 0.1 and a maximum of 8 in each graph for display purposes.

### Figure arrangement

Final figures were arranged in Adobe Illustrator (26.0.1).

## Data Availability

All data produced in the present study are available upon reasonable request to the authors

## Acknowledgments

The authors would like to thank all participants in the study. We thank and acknowledge Takayuki Shiroyama, Kotaro Miyake, Yasuhiko Suga and Yujiro Naito for contribution to clinical practice. Yoshimi Noda, Takayuki Niitsu, Yuichi Adachi, Takatoshi Enomoto, Saori Amiya, Reina Hara, Makoto Yamamoto, Tomoki Kuge, Kinnosuke Matsumoto, Midori Yoneda, Yuji Yamamoto, Yuko Yoshimine, Saki Minoda, Takehiro Hirayama, Kenji Funakoshi, Yasutaka Okita, Shoji Kawada, Daisuke Nakatsubo, Tomomi Tada, Masashi Okamoto, and Hiroshi Shimagami for contribution to clinical practice and sample collection. Ayako Takuwa for sample transport. Rika Ishii for Technical support. Shuhei Sakikabara, Yuchen Liu, Fuminori Sugihara, David Priest, Kelvin Chen, and Yusuke Takeshima for helpful discussions.

## Funding

Japan Society for the Promotion of Science, Kakenhi 19H01021, 20K21834 (YO)

Japan Agency for Medical Research and Development: JP21km0405211, JP21ek0109413, JP21ek0410075, JP21gm4010006, and JP21km0405217 (YO)

JST Moonshot R&D: JPMJMS2021, JPMJMS2024 (YO)

Takeda Science Foundation: (YO)

Bioinformatics Initiative of Osaka University Graduate School of Medicine, Osaka University: (YO)

IFReC grant program for next generation principal investigators (JBW)

Japan Society for the Promotion of Science 16H06295 (SS)

Japan Agency for Medical Research and Development JP19gm0010005 (SS)

Leading Advanced Projects for Medical Innovation (SS)

## Author contributions

Conceptualization: YO, AK, JBW

Methodology: YO, RE, YS, YY, TM, TM (T. Morita), YK, AK, JBW

Investigation: JBW, JT, RI

Data curation: RE, YS, YO, YY, TM, TM (T. Morita), YK

Visualization: JBW, JNS

Funding acquisition: YO, JBW, SS

Project administration: YO, AK, JBW, HH, YT

Resources: YY, TM, TM (T. Morita), YK, HH, YT, RE, YS, YO, DO

Supervision: YO, AK, JBW, SS, DO

Writing – original draft: JBW

Writing – review & editing: JBW, YO, SS, JNS, DP, JT.

## Data and materials availability

Mass cytometry data is uploaded to flow repository ID: FR-FCM-Z4XN and will be made publicly available upon publication. R code is available on request.

## Extended data

**Extended data Fig.1.**
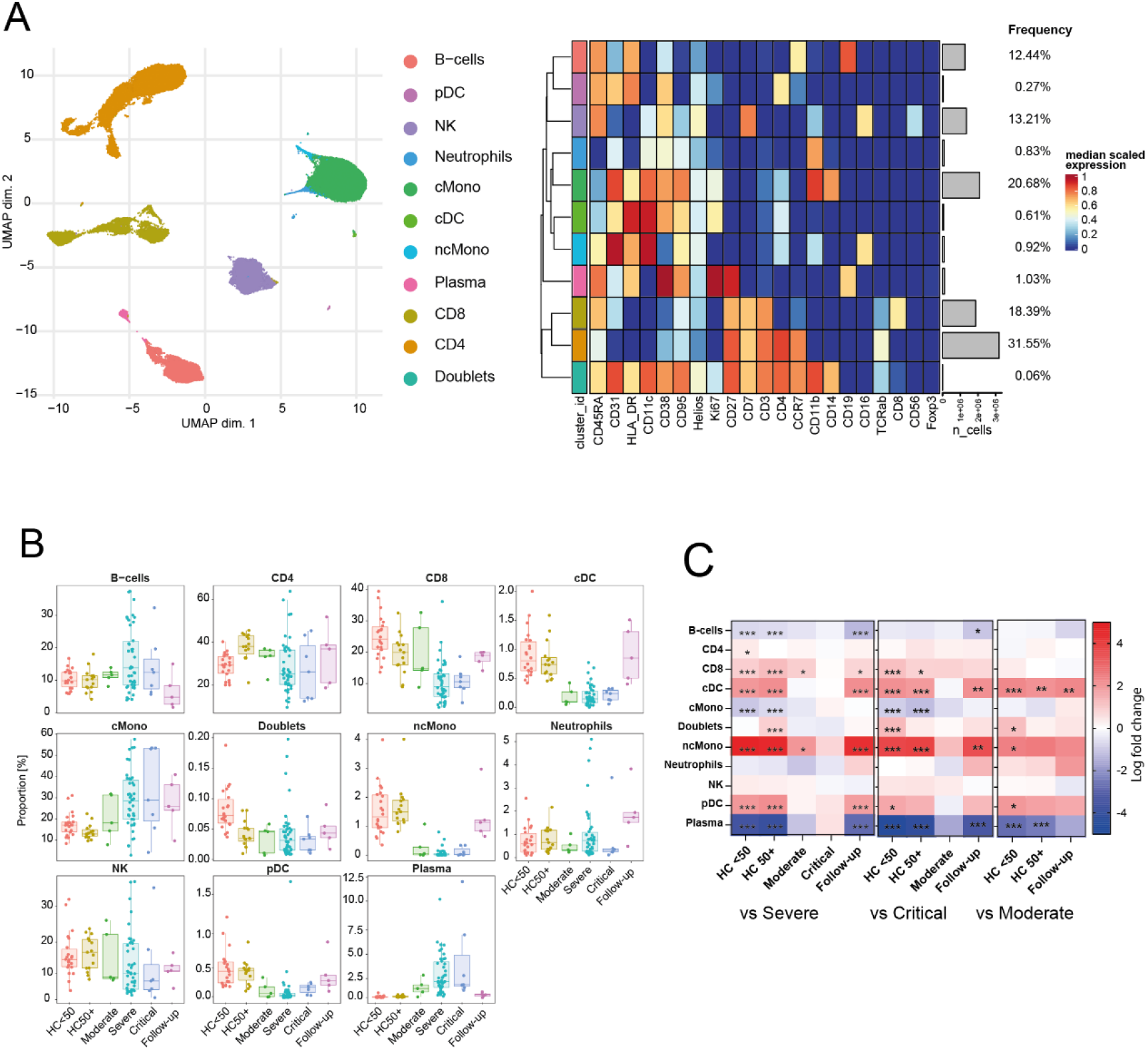
Alterations to frequency of CD45^+^ cells. **A)** UMAP and expression heatmap of FlowSOM clusters from 10,000,000 cells across 100 donors. **B)** Frequency boxplots of proportion of CD45^+^ cell from indicated clusters. Healthy controls under 50 years of age (HC<50), healthy control of 50 or over (HC50+), moderate, severe, critical or follow-up COVID-19 patients. **C)** Comparison of the fold change (log_2_) in cluster-frequency between the indicated group and severe (left), critical (middle), and moderate (right) COVID-19 patients. *p≤0.05, **p≤0.01, ***p≤0.001 by edgeR.

**Extended data Fig.2.**
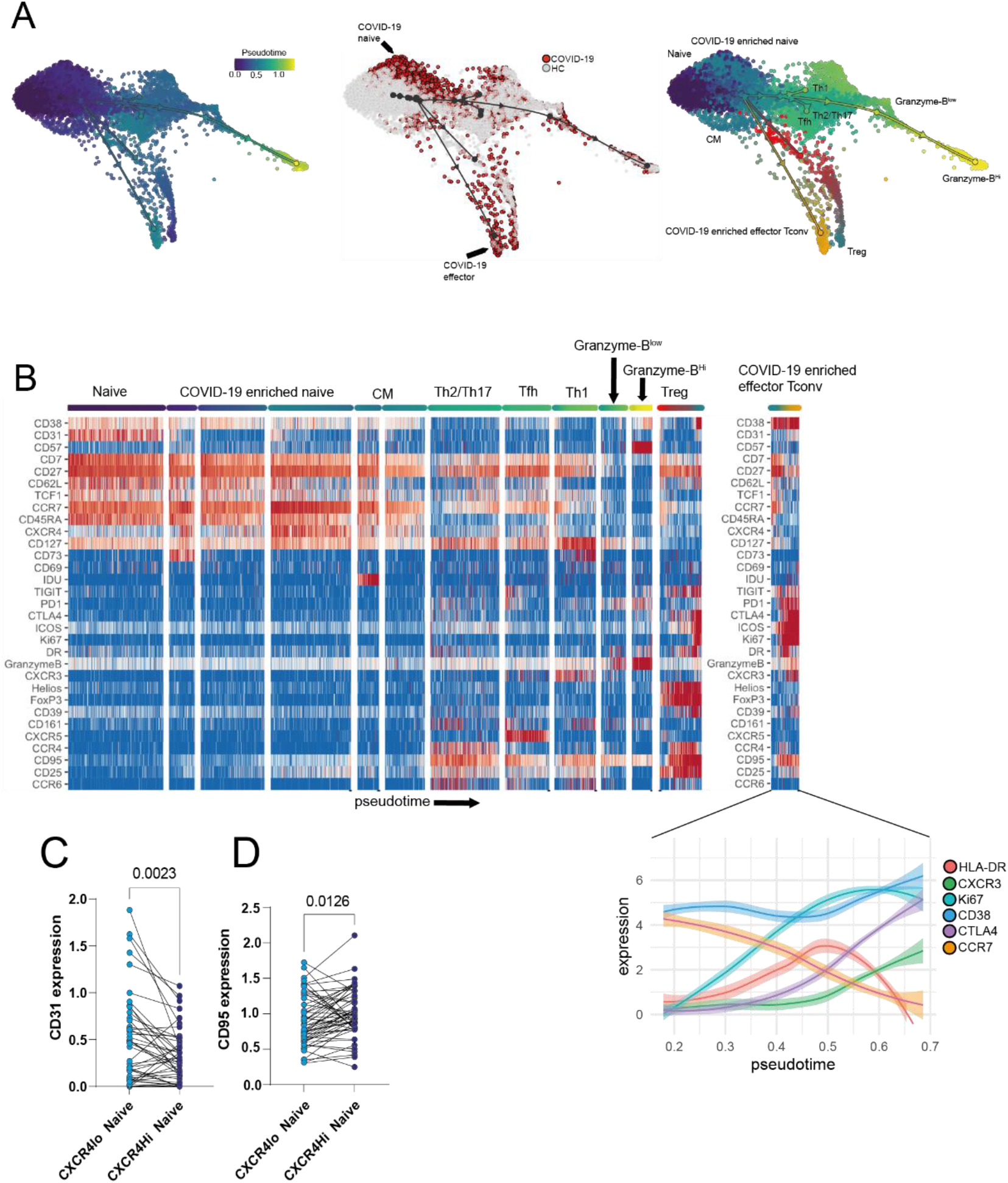
Trajectory analysis of CD4 T-cells in COVID-19. **A)** Force-Atlas2 visualization of CD4 T-cell pseudo time (left) and identified milestones corresponding to specific T cell subsets (right). The middle figure indicates single cells originating from COVID-19 patients or healthy controls. **B)** Heatmap of marker expressions within milestones/subsets. **C)** CD31 expression within clusters CXCR4^lo^ naïve and CXCR4^hi^ naïve from Fig. 2A. **D)** CD95 expression within clusters CXCR4^lo^ naïve and CXCR4^hi^ naïve from Fig. 2A. Central memory (CM).

**Extended data Fig.3.**
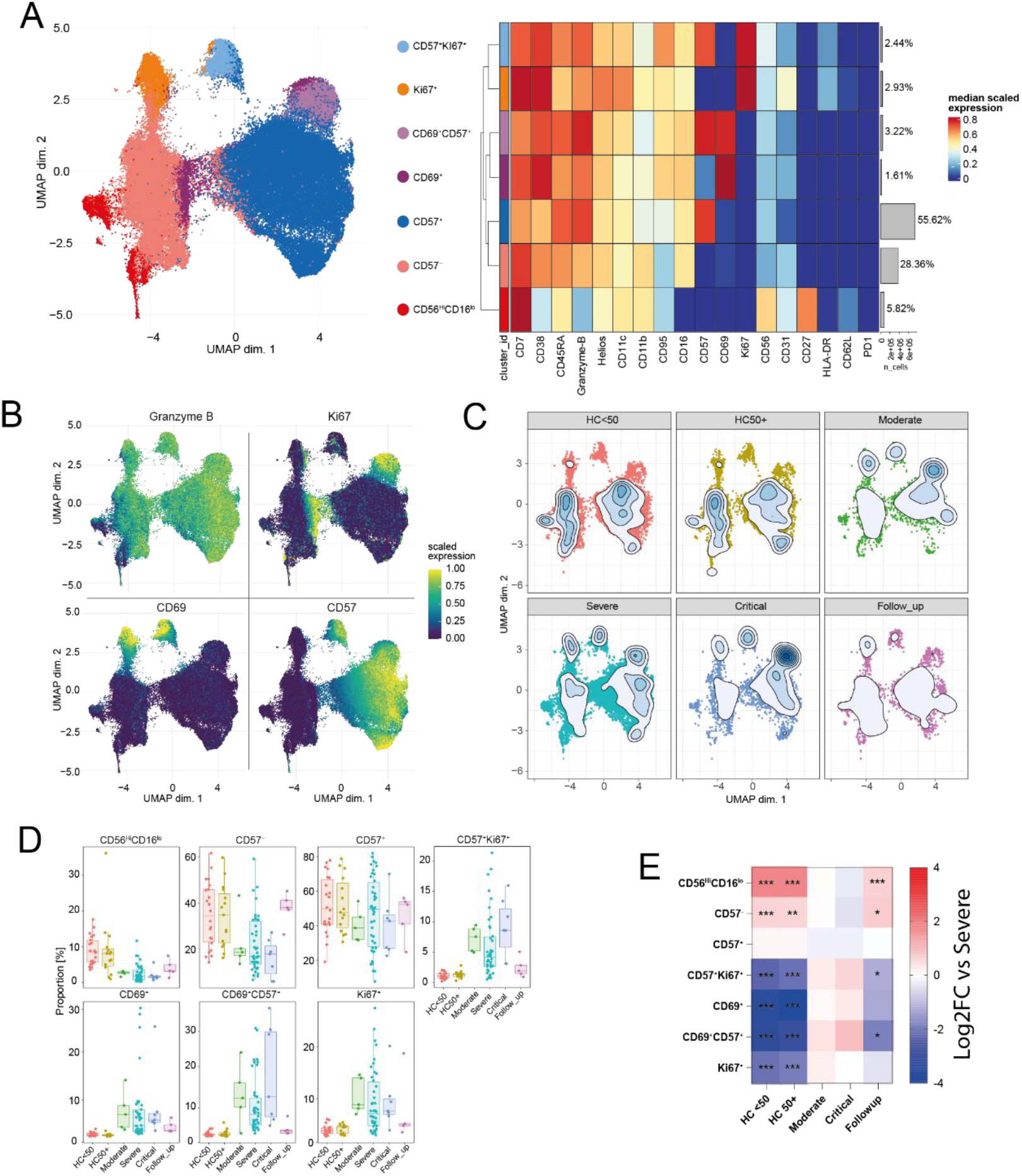
NK phenotypes in COVID-19. **A**) UMAP and expression heatmaps of annotated FlowSOM clusters from NK cells (cluster NK from Extended data Fig.1A). **B)** Scaled expression of indicated markers displayed on the UMAP. **C)** Population density of cells displayed on UMAP. **D)** Frequency boxplots of proportion of CD45^+^ from indicated clusters. Healthy controls under 50 years of age (HC<50), healthy control of 50 or over (HC50+), moderate, severe, critical or follow-up COVID-19 patients. **E)** Comparison of the fold change (log_2_) in cluster-frequency between the indicated group and severe COVID-19 patients. *p≤0.05, **p≤0.01, **p≤0.001 by edgeR.

**Extended data Fig.4.**
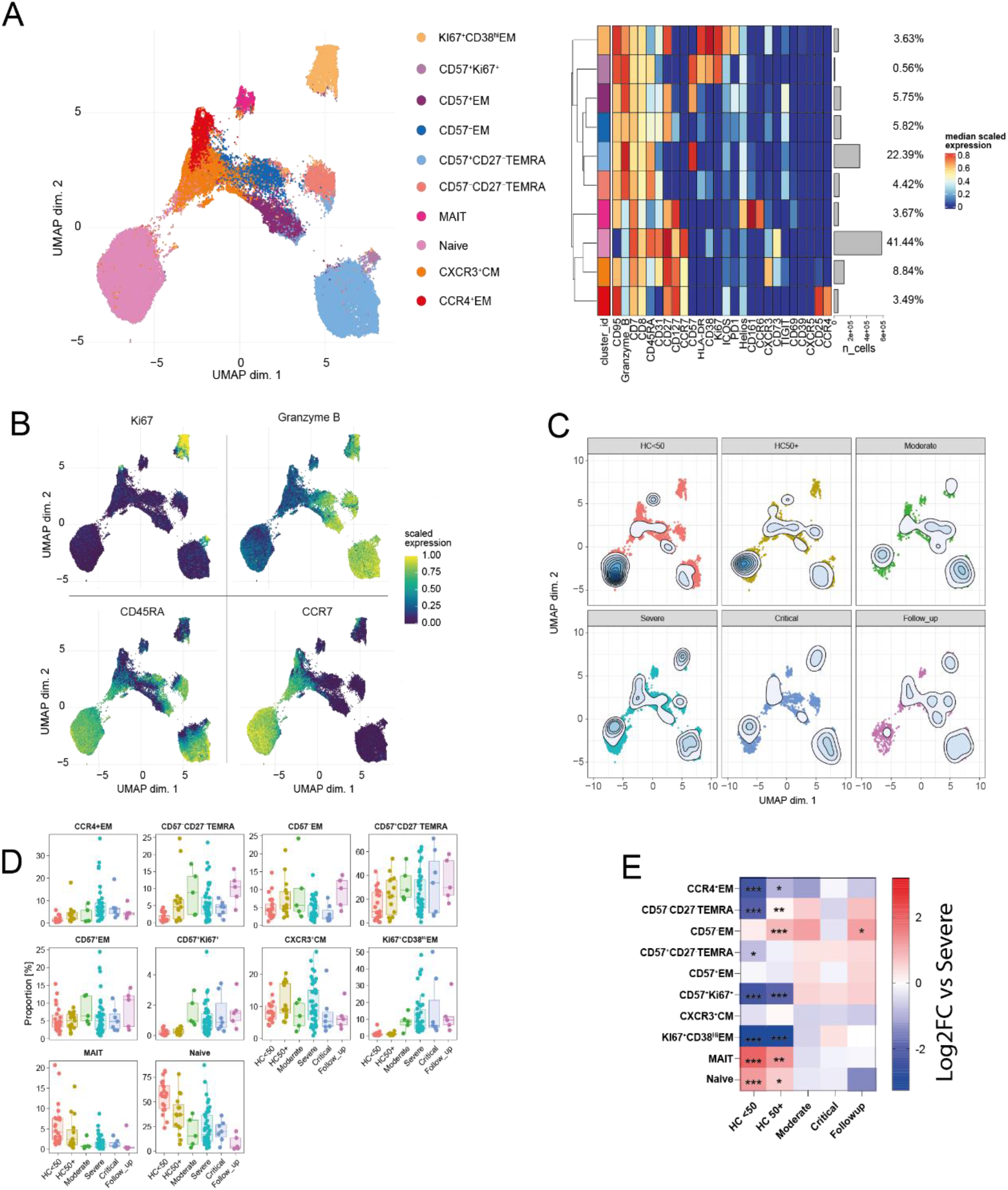
CD8 phenotypes in COVID-19. **A**) UMAP and expression heatmaps of annotated FlowSOM clusters from CD8 cells (cluster CD8 from Extended data Fig.1A). **B)** Scaled expression of indicated markers displayed on the UMAP. **C)** Population density of cells displayed on UMAP. **D)** Frequency boxplots of proportion of CD45^+^ cell from indicated clusters. Healthy controls under 50 years of age (HC<50), healthy control of 50 or over (HC50+), moderate, severe, critical or follow-up COVID-19 patients. **E)** Comparison of the fold change (log_2_) in cluster-frequency between the indicated group and severe COVID-19 patients. *p≤0.05, **p≤0.01, **p≤0.001 by edgeR. Effector memory (EM), Central memory (CM), Terminal effector CD45RA positive (TEMRA), Mucosal associated invariant T (MAIT).

**Extended data Fig.5.**
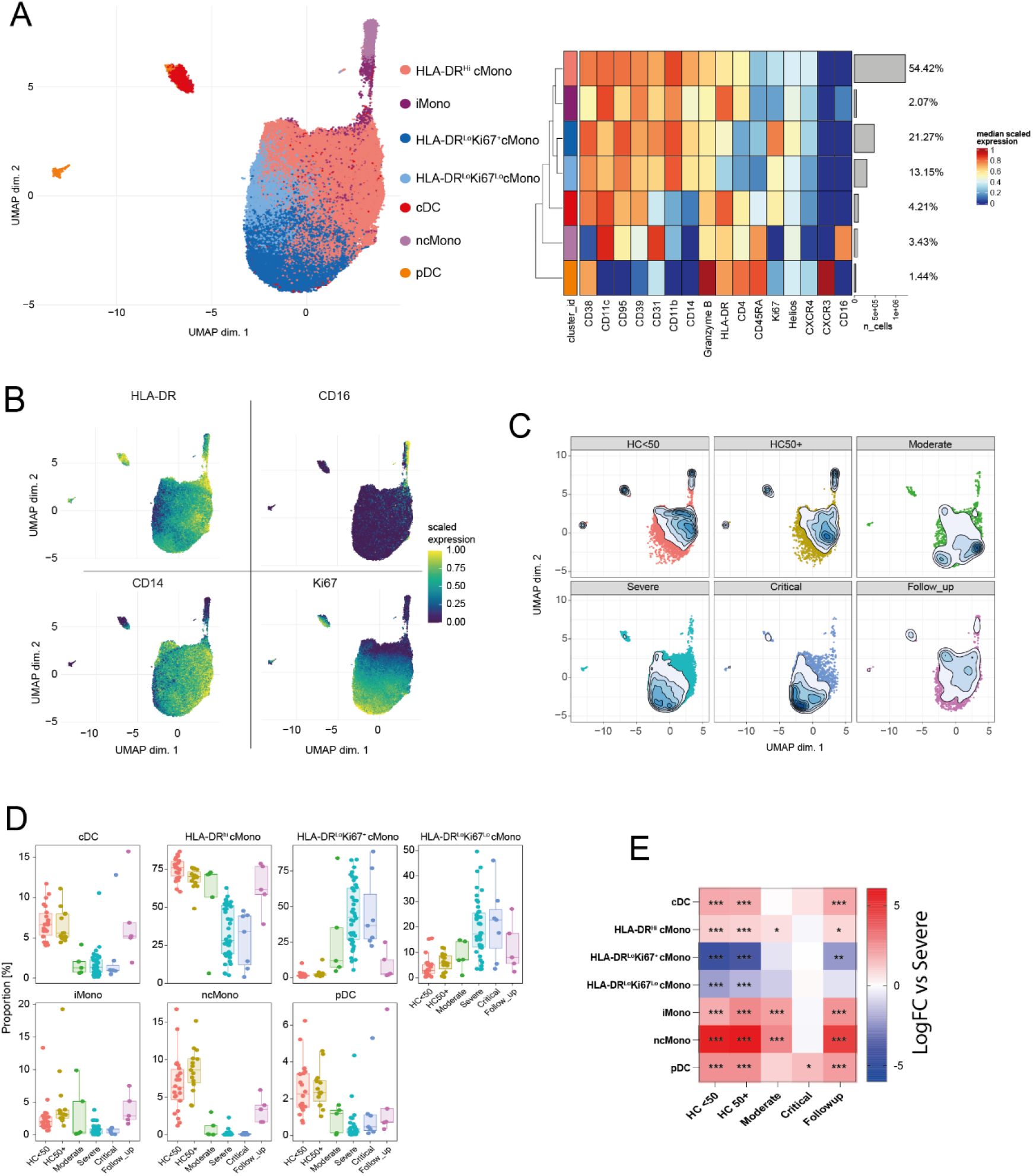
Myeloid cell and DC phenotypes in COVID-19. **A**) UMAP and expression heatmaps of annotated FlowSOM clusters from myeloid cells and DCs (clusters cDC, pDC, cMono and ncMono from Extended data Fig.1A) **B)** Scaled expression of indicated markers displayed on the UMAP. **C)** Population density of cells displayed on UMAP. **D)** Frequency boxplots of proportion of CD45 ^+^ cell from indicated clusters. Healthy controls under 50 years of age (HC<50), healthy control of 50 or over (HC50+), moderate, severe, critical or follow-up COVID-19 patients. **E)** Comparison of the fold change (log_2_) in cluster-frequency between the indicated group and severe COVID-19 patients. *p≤0.05, **p≤0.01, **p≤0.001 by edgeR.

**Extended data Fig.6.**
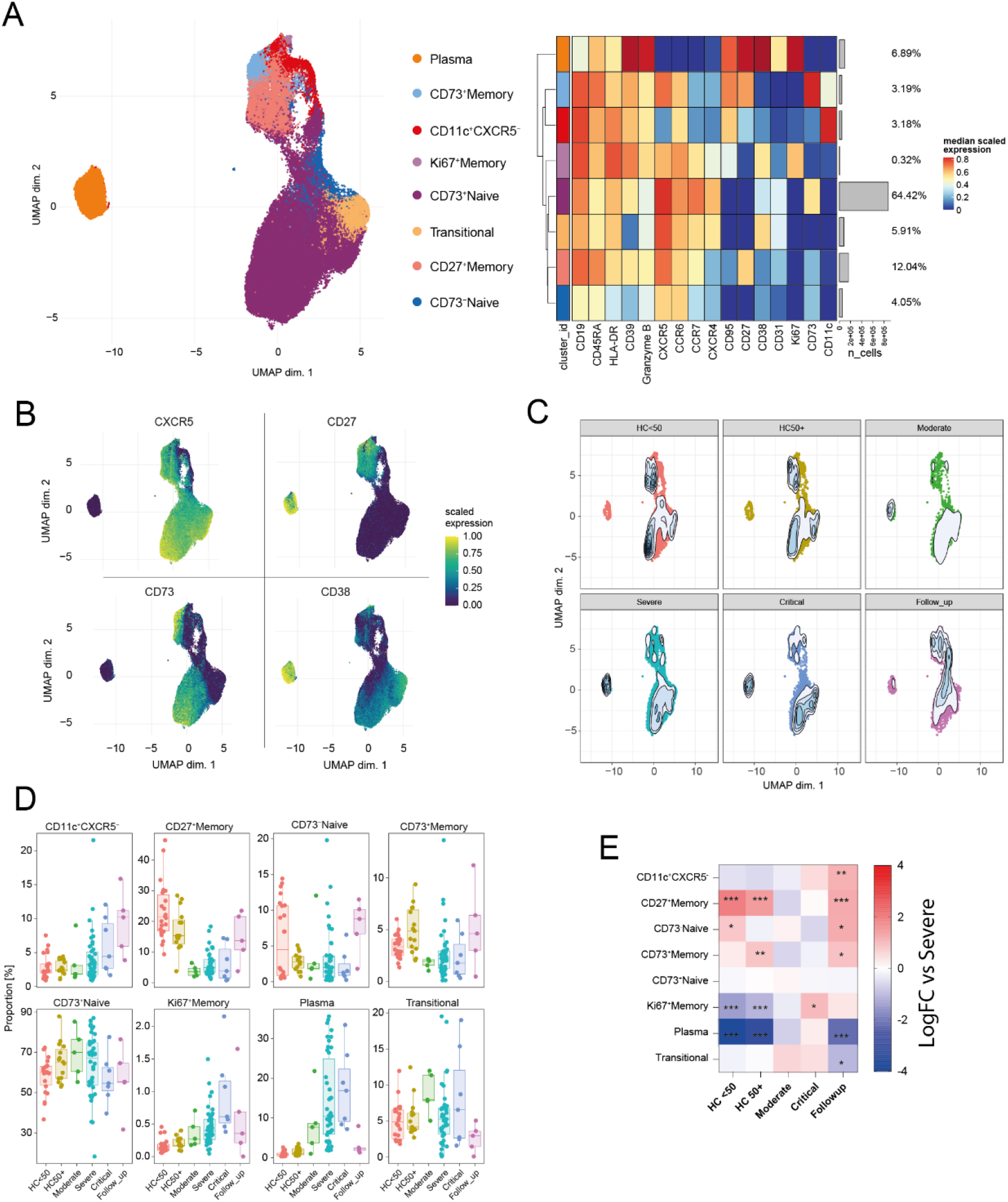
B-cell and plasma cell phenotypes in COVID-19. **A**) UMAP and expression heatmaps of annotated FlowSOM clusters from B and plasma cells (clusters “B-cells” and “plasma cells” from Extended data Fig.1A). **B)** Scaled expression of indicated markers displayed on the UMAP. **C)** Population density of cells displayed on UMAP. **D)** Frequency boxplots of proportion of CD45^+^ cell from indicated clusters. Healthy controls under 50 years of age (HC<50), healthy control of 50 or over (HC50+), moderate, severe, critical or follow-up COVID-19 patients. **E)** Comparison of the fold change (log2) in cluster-frequency between the indicated group and severe COVID-19 patients. *p≤0.05, **p≤0.01, **p≤0.001 by edgeR.

**Extended data Fig.7.**
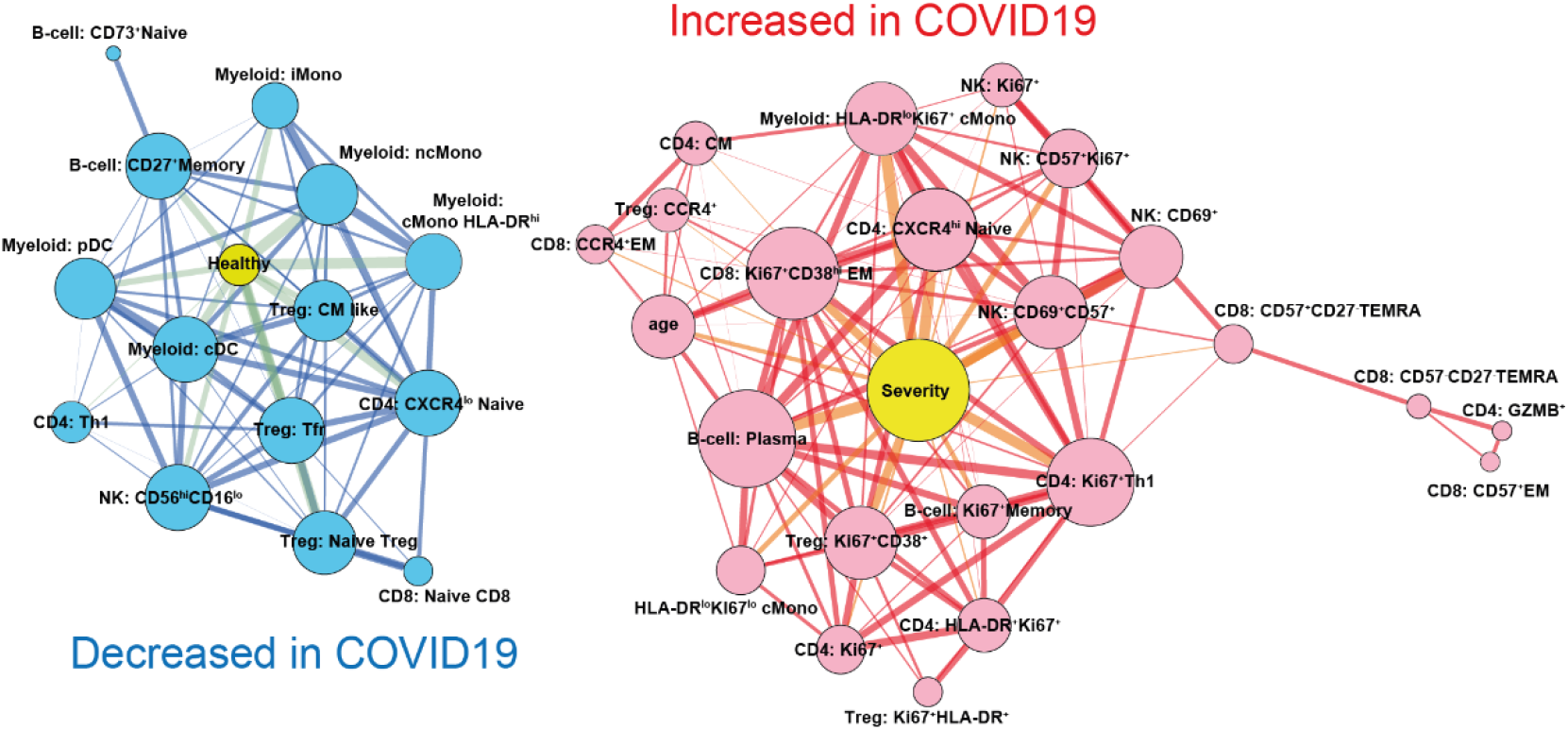
Broad changes to cellular networks in COVID-19. Spearman correlation network of cellular subset frequencies between indicated subsets, COVID score (0 = healthy, 1 = COVID-19 patient) and sex (1 = male, 2= female). Edge width is proportional to correlation strength, node size is proportional to the number of connecting edges. Edges connecting to other cellular subsets are positive correlations between indicated subsets. Layout by ForceAtlas2 using edge weights as input.

**Extended data Fig. 8.**
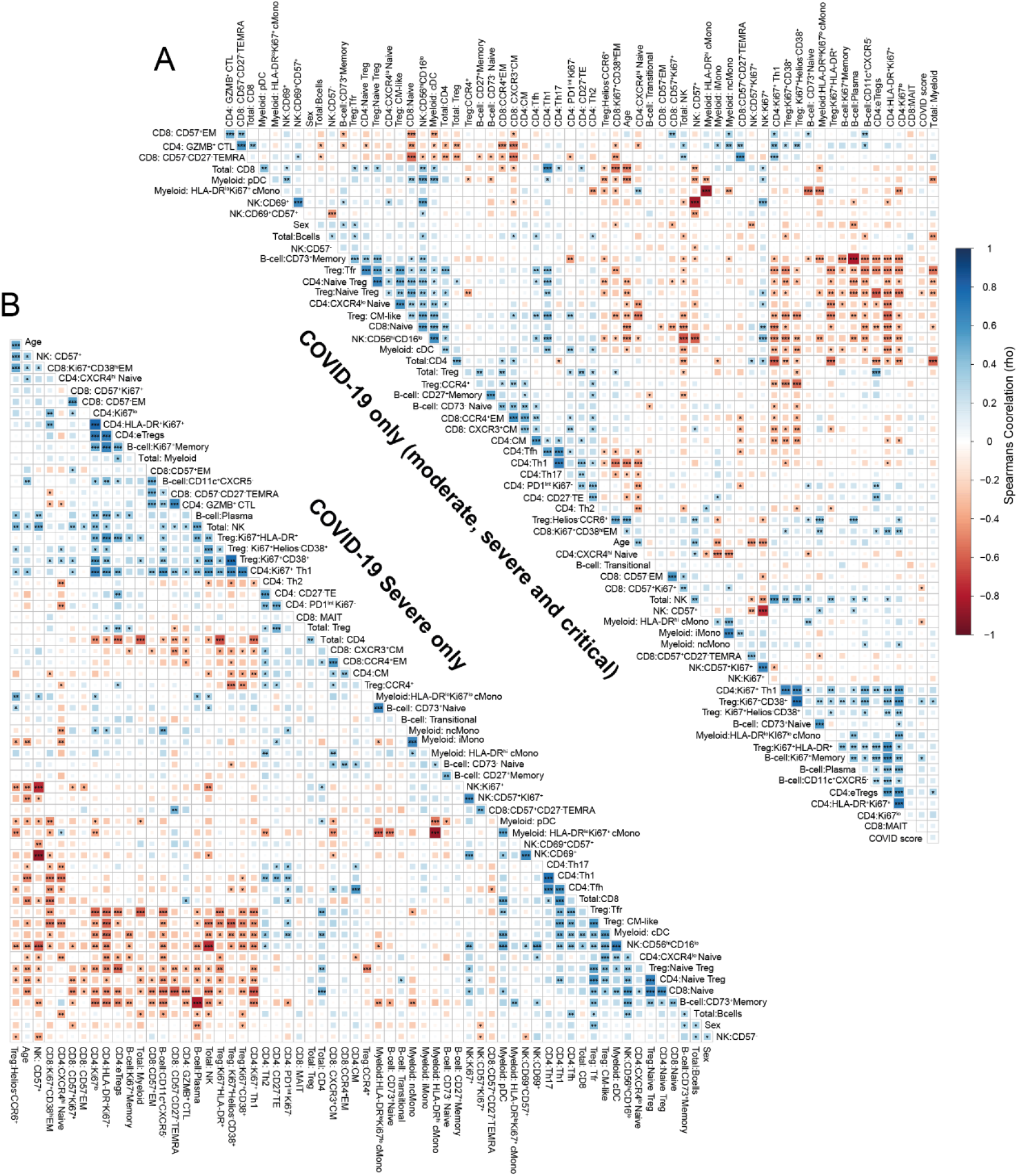
Cellular correlations in patient subgroups. **A)** Spearman correlations matrix of subset frequencies from moderate, severe, and critical patient groups with severity of infection scored as WHO ordinal scale (4 = moderate, 6= severe, 7 = Critical) and sex (1 = male, 2= female). **B)** Spearman correlations matrix of subset frequencies from severe COVID-19 patients with severity of infection scored as WHO ordinal scale (4 = moderate, 6= severe, 7 = Critical) and sex (1 = male, 2= female) Correlations by spearman rank (A, B, C). Significance *p≤0.05, **p≤0.01, ***p≤0.001. Effector memory (EM), Central memory (CM), Terminal effector CD45RA positive (TEMRA).

**Table S1.**
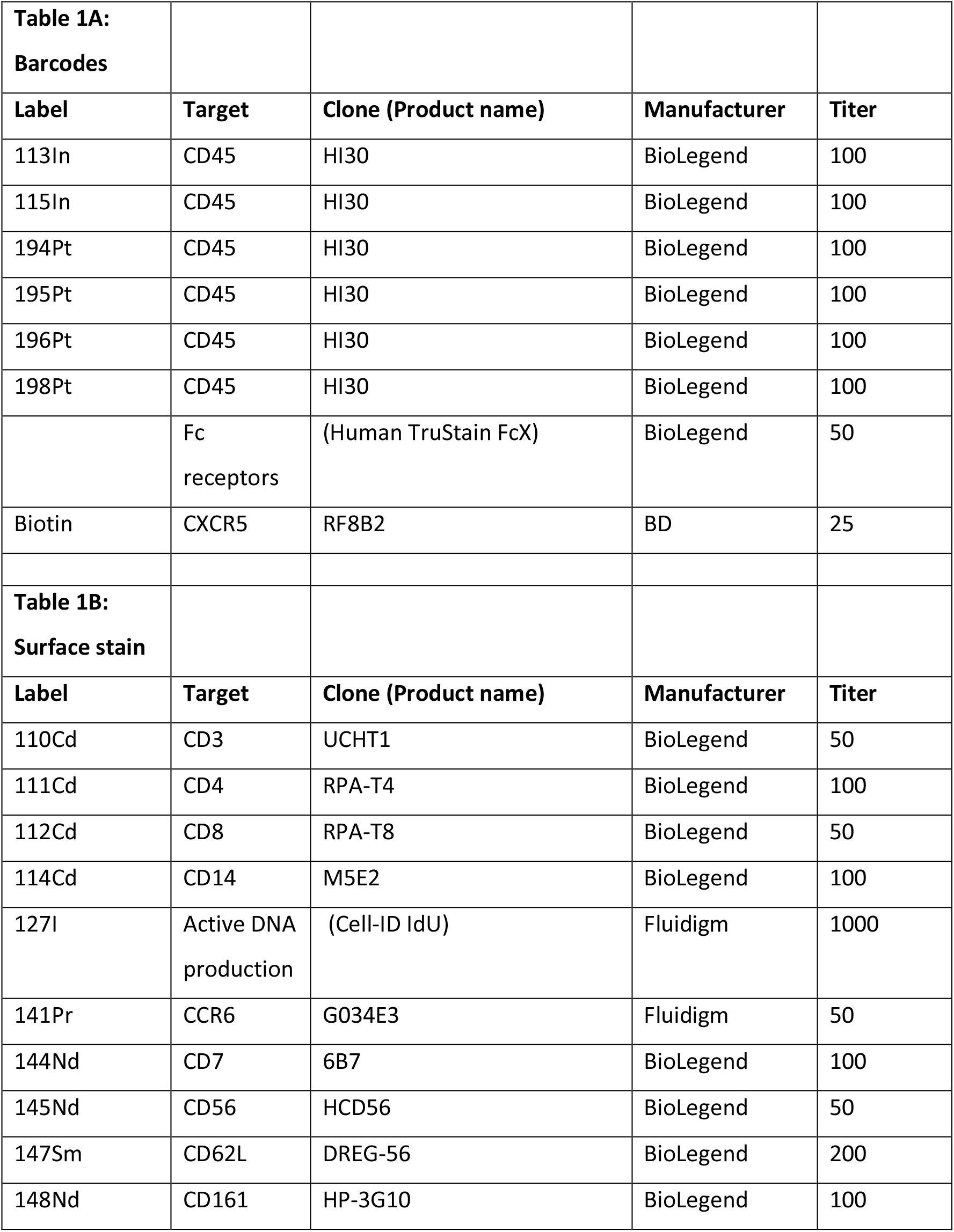
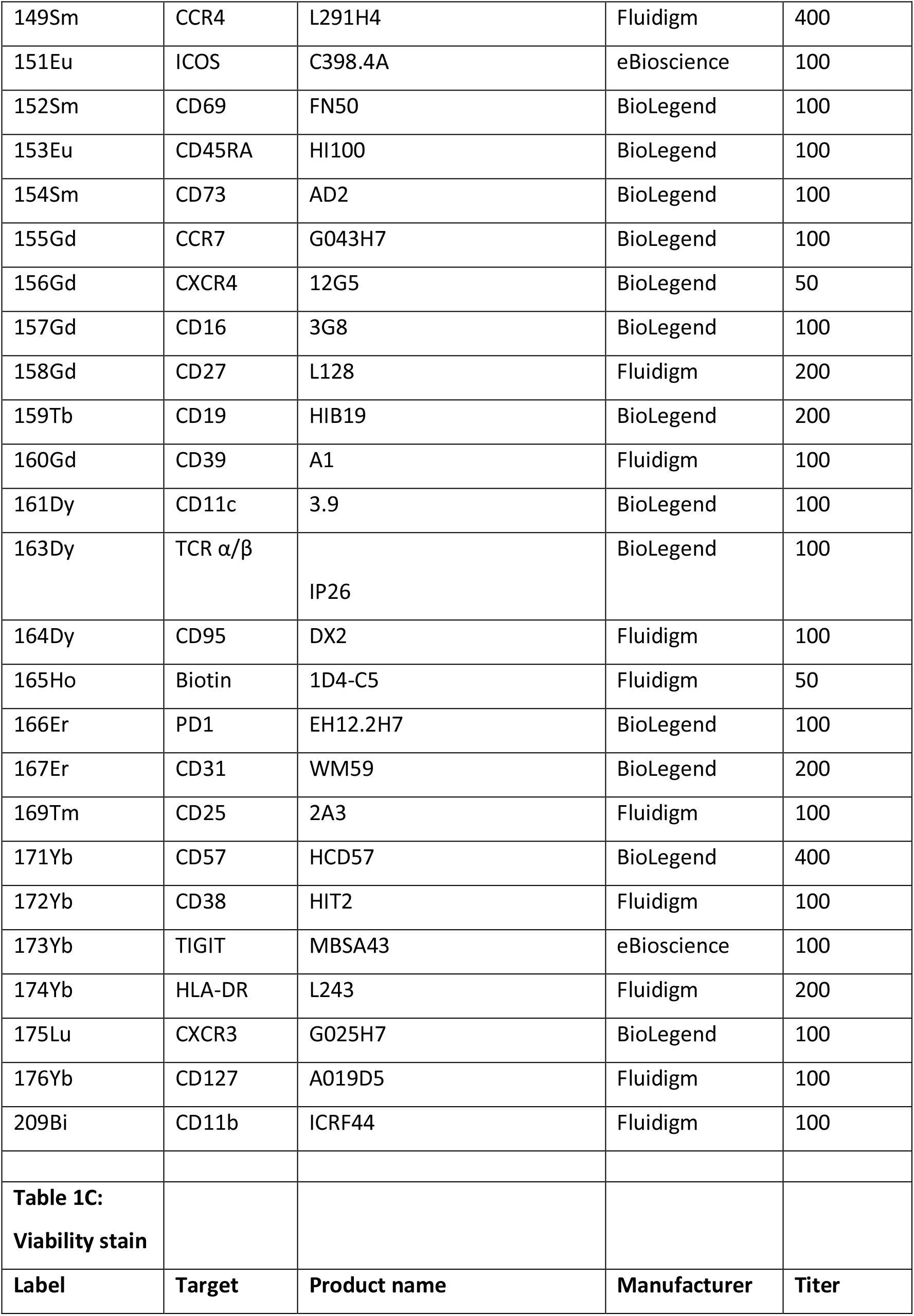
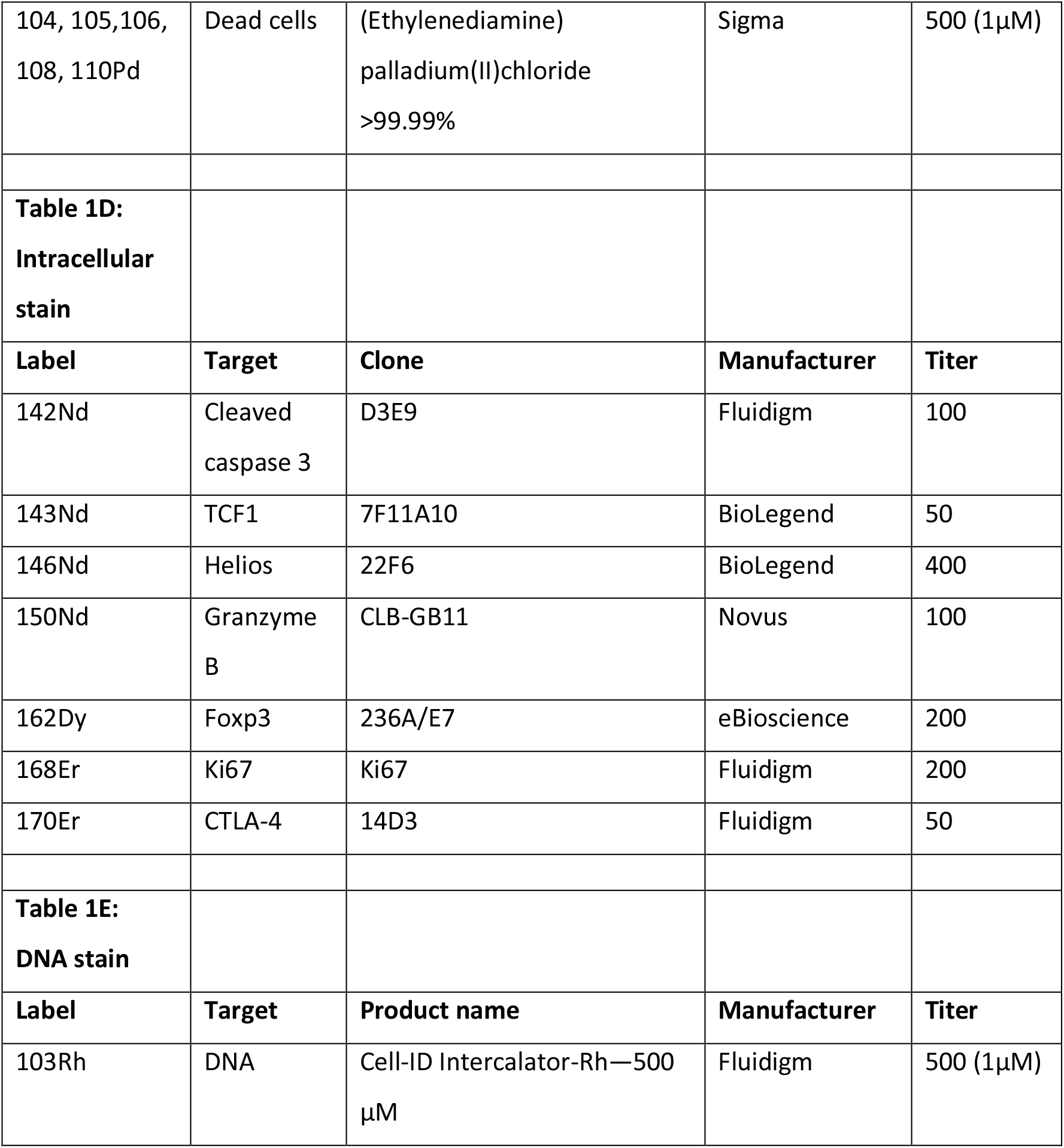
Mass cytometry staining panels.

